# Human aging reflects increases in entropy across organ networks

**DOI:** 10.1101/2025.05.06.25327108

**Authors:** Meng Hao, Hui Zhang, Yi Li, Yaqi Huang, Jingyi Wu, Shuishan Zhang, Xianglin Aneta Guo, Xueqin Li, Zixin Hu, Xiangnan Li, Shuai Jiang, Kamaryn T. Tanner, Andrew Rutenberg, Jie Chen, Zhijun Bao, Jiucun Wang, Yiqin Huang, Alan A Cohen, Li Jin, Xiaofeng Wang

**Affiliations:** Shanghai Key Laboratory of Clinical Geriatric Medicine, Shanghai Institute of Geriatrics and Gerontology, Huadong Hospital, Shanghai Medical College, Fudan University, Shanghai, 200040, China; Human Phenome Institute, Zhangjiang Fudan International Innovation Centre, Fudan University, Shanghai, 201203, China; Fudan Zhangjiang Institute, Shanghai 201203, China; School of Global Health, Chinese Center for Tropical Diseases Research, Shanghai Jiao Tong University School of Medicine, Shanghai, 200025, China; Department of Vascular Surgery, Shanghai Key Laboratory of Vascular Lesion Regulation and Remodeling, Shanghai Pudong Hospital, Fudan University Pudong Medical Center, Shanghai, 201399, China; Butler Columbia Aging Center, Mailman School of Public Health, Columbia University, NY 10032, America; Department of Physics and Atmospheric Science, Dalhousie University, Halifax, NS B3H 4R2, Canada; Department of Environmental Health Sciences, Mailman School of Public Health, Columbia University, NY 10032, America

## Abstract

Aging involves diminished homeostatic control and changes in individual biomarker levels/states. However, it is unknown whether these alterations reflect a rise in entropy during the aging process, and whether entropy disrupts broad systemic interrelationships^1–4^. The entropy of human aging has not been well characterized, but measures of systemic entropy could reveal aging dynamics that may not be apparent even by integration of state-based aging metrics^5–7^. Here, we leverage the Distance of Covariance (DISCO), which quantifies entropy in large ensembles of biological information, to demonstrate that organs and systems exhibit interconnected increased entropy with age. We validate DISCO on multiple data substrates (clinical biomarkers, proteomics, metabolomics, and microbiomes) in five cohort datasets: UK BioBank, National Health and Nutrition Examination Survey, and three Chinese cohorts of older adults. DISCO consistently outperforms mortality prediction of existing metrics of dysregulation and is comparable to the best-in-class epigenetic clocks. It also strongly predicts frailty and incidence of age-related chronic conditions. Crucially, organ- and system-specific DISCO scores derived from circulating proteomics demonstrate broad predictive power with little to no specificity of a given organ predicting its own diseases and mortality. Network analysis of organ- and system-specific DISCO shows that more central, connected organ DISCOs predict health outcomes more strongly. For example, for each mortality cause, brain entropy is one of the strongest predictors. These findings challenge current notions of independent organ-specific aging signatures^8–10^, suggesting instead that while pathology may be organ-specific, entropy spills readily across systems, and thus conversely that health during aging requires integrated homeostatic coordination across multiple systems.

---

Despite decades of research, there is no consensus either on the nature of the aging process^11,12^ or on how to measure it^13,14^. Recently, there has been substantial interest in the hypothesis that the aging process is a manifestation of increasing entropy in biological systems, with this rise in disorder at the core of its progression^6,7,15,16^. Under this hypothesis, organisms are complex information systems that resist the tendency toward entropy under the second law of thermodynamics only via inputs of energy from their environment and via sophisticated regulatory mechanisms that prevent and repair damage or other forms of information loss. Because these mechanisms are imperfect, damage nevertheless accrues, and entropy increases over time, causing a vicious cycle and accelerating the aging process. In parallel, a related hypothesis posits that aging does not occur independently cell by cell, tissue by tissue, or organ by organ, but rather represents a breakdown in the coordination across systems due to feedback effects as entropy accrues in individual systems^4^.

However, tests of these hypotheses have been hindered by the lack of appropriate measurement tools for entropy. Shannon entropy is measured based on a distribution and is thus not readily applied to a single data point for an individual, even if the data point is a high-dimensional-omics profile^7^. Homeostatic dysregulation metrics present a possible solution to measurement of entropy, in which homeostatic dysregulation quantify an organism’s deviation from physiological norms, a state expected to increase as systemic entropy rises. Recent work using ECG-derived entropy based on homeostatic dysregulation has shown predictive value for mortality and fractures. However, a direct method for quantifying entropy from individual high-dimensional data (e.g., omics profiles) is needed.

Here, we introduce the DIStance of COvariance (DISCO), a newly developed approach to measure entropy in individuals. DISCO quantifies how much any individual sample perturbs the correlation structure of the ensemble when it is added or removed (**Fig. 1a**). A sample that minimally perturbs the ensemble adds little new information and is thus considered closer to the homeostatic norm of the population; conversely, a sample that perturbs it heavily is outside the norms and is considered more dysregulated. We validated DISCO as a metric of entropy and then applied it to test whether entropy would reveal more interconnected signals of aging than traditional organ aging “clocks.”

**Figure 1.**
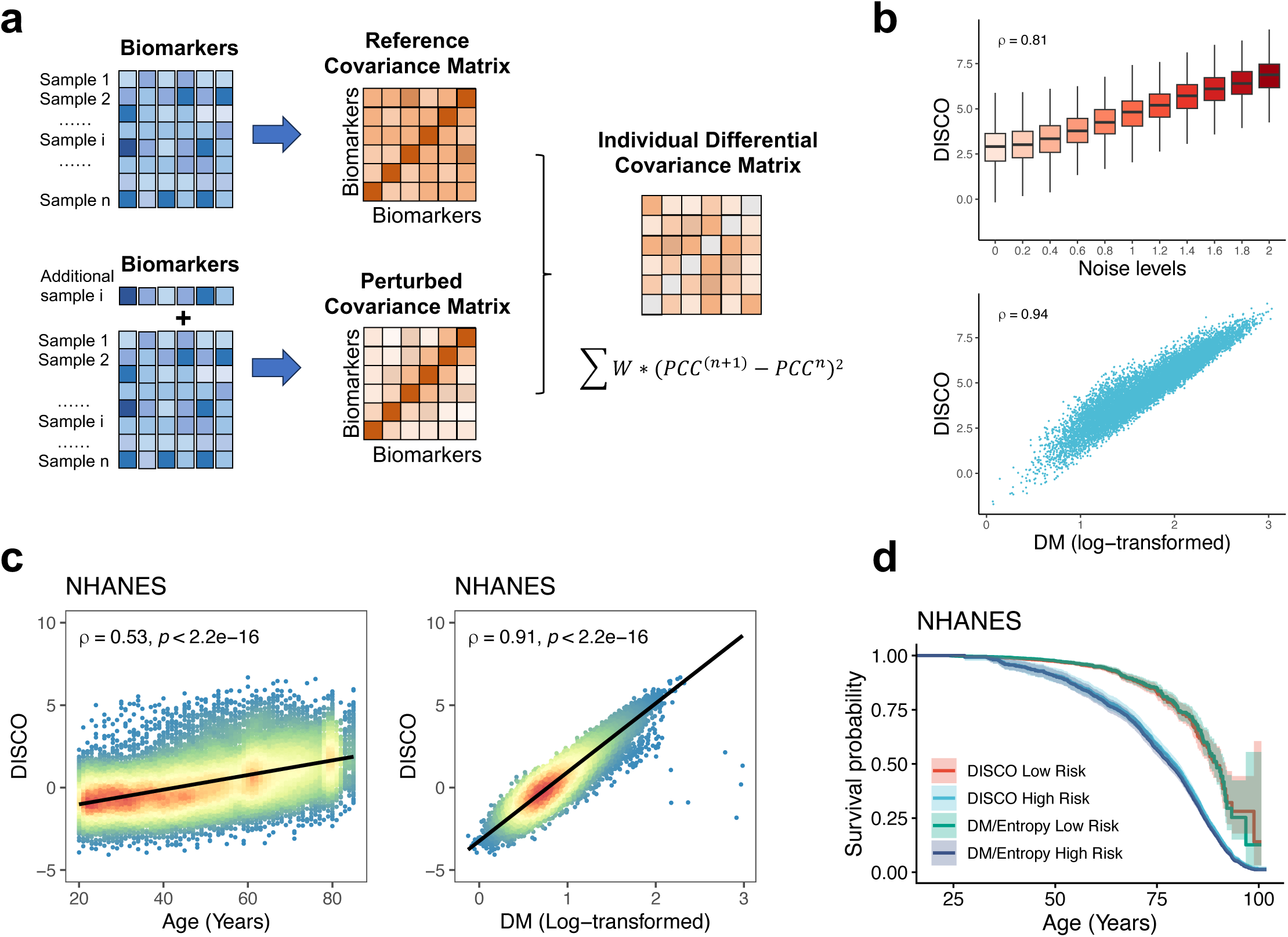
The DISCO framework, simulation analysis and its application to clinical biomarkers. (**a**) Conceptual basis of distance of covariance (DISCO): The phenotypic covariance matrix changes slightly with an additional sample (e.g, i^th^), highlighting DISCO as an individual-level metric quantifying aggregated covariance difference. Boxplot and Scatter plots (**b**) of DISCO and Mahalanobis distance (DM) with increased noise to Gaussian distributed datasets with simulations. (**c**) DISCO versus age (left) and DM (right) in the NHANES cohort, with Spearman’s correlation coefficients (ρ) and p-values. Kaplan-Meier curves (**d**) compared survival probabilities for individuals categorized by low/high DISCO and DM/Entropy risks (first and forth quartiles) in NHANES, respectively, using chronological age as the time-scale axis. The DM and Entropy risks are identical, represented by the same line.

We first tested the hypothesis that DISCO quantifies entropy via homeostatic dysregulation, benchmarking it against the existing Mahalanobis distance (DM) approach. Mathematically, DISCO and DM are related to Shannon and related metrics of entropy, operationalizing them on an individual scale (**Supplementary Materials**). Importantly, we prove that the entropy distance for an individual, the entropy shift induced by perturbing that individual into a reference population, is mathematically equivalent to DM, a result validated in simulations and real data (Extended Data Fig. 1-2). In simulated scenarios of increasing entropy (**Fig. 1b**), DISCO levels rose monotonically (Spearman’s ρ = 0.81) and showed a strong correlation with DM (ρ = 0.94). When applied to clinical biomarker data, DISCO was strongly correlated with DM in five independent cohort datasets of aging (National Health and Nutrition Examination Survey [NHANES], UK Biobank [UKB], the China Health and Retirement Longitudinal Study [CHARLS], the Rugao Longitudinal Aging Study [RuLAS], and the Chinese Longitudinal Healthy Longevity Survey [CLHLS]; 0.7< ρ < 0.91, p < 0.0001 for all, Extended Data Fig. 3). Like DM, DISCO was also weakly to moderately associated with age cross-sectionally (0.2 < ρ < 0.53, p < 0.0001 for all). In all cases, compared with DM, DISCO also clearly distinguished mortality risk (hazard ratio per unit after age- and sex- adjustment (aHR): 1.26 < aHR < 1.71, p < 0.0001, [concordance index (C-indices), from 0.65 to 0.77], Extended Data Table 1), generally with a magnitude similar to DM.

Next, we tested whether DISCO could also be applied in other data modalities, namely proteomics, metabolomics, microbiota, and combinations, including clinical biomarkers. In UKB, we utilized DISCO on blood serum proteomics. Following quality control and feature selection, DISCO analysis included 367 proteins in 47,859 participants. Addition of proteomics substantially strengthened associations with age (ρ = 0.42, etc.) and mortality (C-index of proteomics alone = 0.740, clinical biomarker alone = 0.670, and together = 0.741; **Fig. 2a-b**), clearly demonstrating stronger associations than DM in a way that was not apparent with clinical biomarkers alone. These results were validated through sensitivity analysis across selections of biomarkers, reference samples, and distance metrics (Extended Data Fig. 4) and simulation analysis (Extended Data Fig. 5). Using Wave 4 of the RuLAS study (n = 2120), we tested two-year mortality prediction of metabolomics, microbiota, and aggregate (clinical + metabolomics + microbiota) versions of DISCO (**Fig. 2c-f**). Metabolomic entropy, as measured by DISCO analysis on NMR data, was associated with an increased risk of two-year mortality (aHR: 1.17, 95% CI: 1.06-1.29, C-index: 0.578). We then identified 27 microbial species correlated with host metabolism entropy (Spearman p < 0.05) and constructed a DISCO score for the microbiota. Gut microbial entropy demonstrated a stronger association with two-year mortality risk (aHR: 1.38, 95% CI: 1.10-1.74, C-index: 0.610). When integrating microbiota and metabolic entropy with clinical biomarkers, the aggregate DISCO score significantly predicted mortality (aHR: 2.49, 95% CI: 1.92-3.24, C-index: 0.713), surpassing the predictive power of any single data type DISCO.

**Fig. 2.**
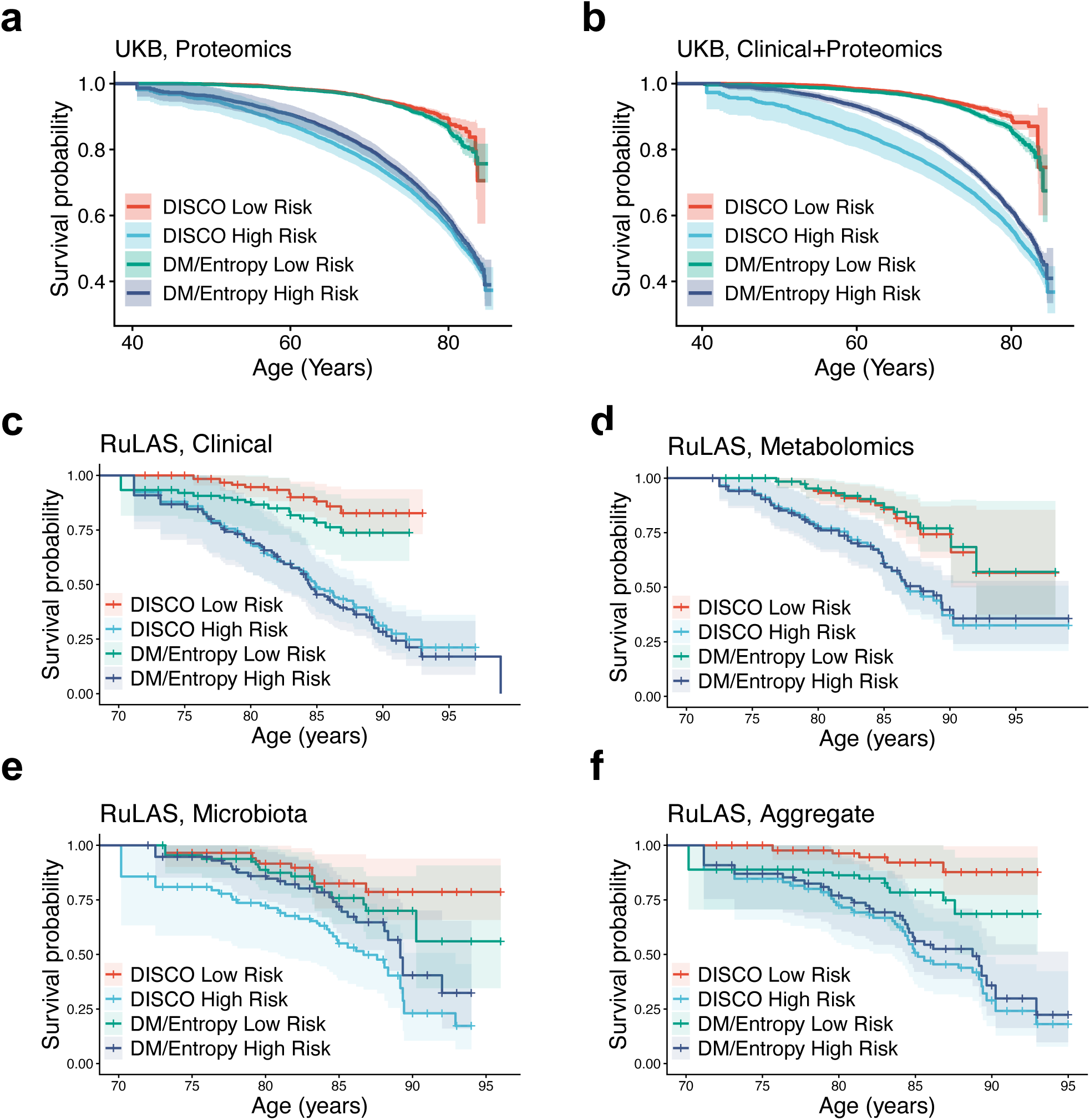
DISCO applications across diverse omics biomarker domains. The Kaplan-Meier (KM) curves of DISCO in UKB using proteomics (**a, b**) and union sets of proteomics and multi-omics (**c-f**) in the RulAS. The individuals stratified by the DISCO and DM/Entropy low and high risks (the first and fourth quartiles) with age as the time-scale axis. The RuLAS cohort included clinical, metabolomic, gut microbiota, and aggregate DISCO, and DM/Entropy scores. DM risk and Entropy risk are equivalent measures; thus, their survival curves are superimposed (same colored line).

For mortality prediction benchmarking (**Fig. 3a-b**), DISCO was evaluated against established epigenetic clocks (e.g., HorvathAge^17^, GrimAge^18^) in the NHANES cohort, which included 2,532 participants (mean age 66.13 ± 10.08 years) with DNA methylation profiles. When combined with chronological age, DISCO exhibited predictive accuracy approaching that of GrimAge (C-index: Age+DISCO = 0.751 vs. GrimAge = 0.759; HorvathAge = 0.713). Age-adjusted residual analysis (“Accel”) further validated DISCO’s predictive capability independent of age, ranking second only to GrimAge and its derivatives. Furthermore, because the key epigenetic clocks are more tightly correlated with chronological age than DISCO, DISCO better distinguishes risk across a wide range of ages, as revealed by Kaplan-Meier analysis across age groups (**Fig. 3b**).

**Fig. 3.**
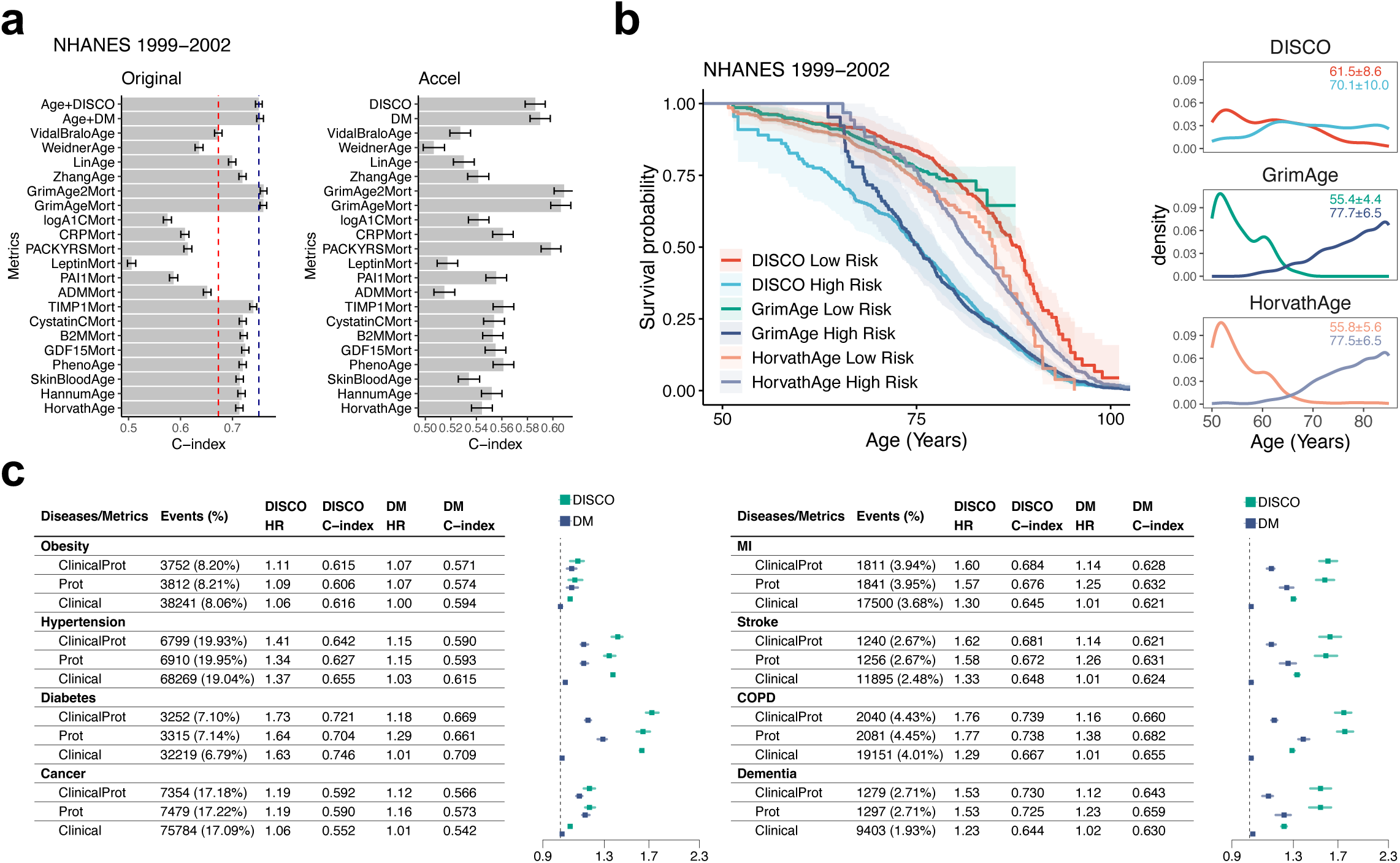
DISCO strongly predicts health outcomes. Bar plots (**a**) compared the discriminative accuracy for mortality (C-index) of DISCO against epigenetic clocks, epigenetic-derived markers, and DM for predicting mortality in NHANES. C-index of DISCO and age were marked with navy blue and red dotted line in the left panel, respectively. The right panel presented the C-index of aging acceleration (Accel), an age-adjusted residual measure. Kaplan-Meier curves (**b**) compared survival probabilities for individuals categorized by low/high DISCO, GrimAge, and HorvathAge risks (first and forth quartiles) in NHANES and UKB, respectively. Right panels: Age distributions (mean ± s.d.) per risk group. Low mortality at younger ages in high-risk GrimAge and HorvathAge clocks in the Kaplan-Meier curves is likely attributable to the small sample at those ages in those groups. Forest plots (**c**) presents hazard ratios (HRs, 95% confidence interval [CI]) for DISCO-associated chronic diseases in the UKB cohort (14-year follow-up). HRs (95% CI) were adjusted for age, sex, BMI, race, education, smoking and drinking status (Model 2). C-index of Model 1 (unadjusted) was shown. DISCO and Mahalanobis distance (DM) were derived from proteomics (Prot), clinical biomarkers, and their combined set (ClinicalProt). Model 1: Disease ∼ DISCO/DM; Model 2: Disease ∼ DISCO/DM + age + sex + covariates. MI: myocardial infarction; COPD: chronic obstructive pulmonary disease.

Next, we tested the relevance of DISCO for predicting clinical outcomes beyond mortality. Using UKB data combining clinical and proteomics markers, we tested associations of DISCO with incidence of eight chronic conditions (obesity, hypertension, diabetes, cancer, myocardial infarction, stroke, COPD, and dementia). In all cases, DISCO was clearly associated with incidence (**Fig. 3c**, Extended Data Fig. 6), with 1.57 < aHR < 2.07 except for cancer, where the association was notably weaker (aHR = 1.11, 95% CI: 1.06-1.15). In all cases, DISCO provided stronger discriminatory power than DM. Likewise, in CHARLS, DISCO was strongly associated with the frailty index (Extended Data Fig. 7); moreover, when DISCO and frailty were used jointly to predict mortality, the DISCO effect dominated.

Given the clear predictive value of broad-spectrum DISCO, we next tested the hypothesis that DISCO of specific organs and tissues would elucidate interconnections during aging. Using the UKB proteomics dataset, we assigned individual proteins to 18 organs/tissues following the methodology of Oh et al.^8^ and calculated DISCO for each set (Extended Data Fig. 8). We then tested the associations of these 18 organ/tissue DISCO scores with incidence of chronic conditions, as well as with all-cause and cause-specific mortality (**Fig. 4a**). In nearly every case, all organ/tissue DISCO scores were positively associated with incidence of all chronic conditions and all causes of mortality; the exception was Alzheimer’s mortality, which was only associated with brain and intestine DISCO scores, concordant with the gut-brain axis^19^. Strikingly, there was little to no specificity of which organ/tissue DISCO scores were associated with which chronic conditions or mortality causes. Some DISCO scores were associated more strongly with outcomes across the board, and some outcomes were more strongly associated with all DISCO scores, but in no case was a single organ/tissue DISCO score associated much more strongly with mortality or disease related to that organ or tissue. Visually, this is apparent in **Fig. 4a** as some rows and some columns being darker red than others, but no cells being markedly darker than all others in their row/column. For example, brain DISCO is among the top predictors of every single mortality cause, and indeed is much more strongly associated with heart and respiratory disease mortality than with Alzheimer’s mortality. Next, we generated scores for each individual in the UKB proteomics dataset for the number of organs/tissues with a score in the top 10%. We plotted the average score by age, which showed an exponential increase nearly perfectly parallel to the increase in mortality with age (**Fig. 4b**), suggesting tight coupling of entropy with both aging and mortality risk.

**Figure 4.**
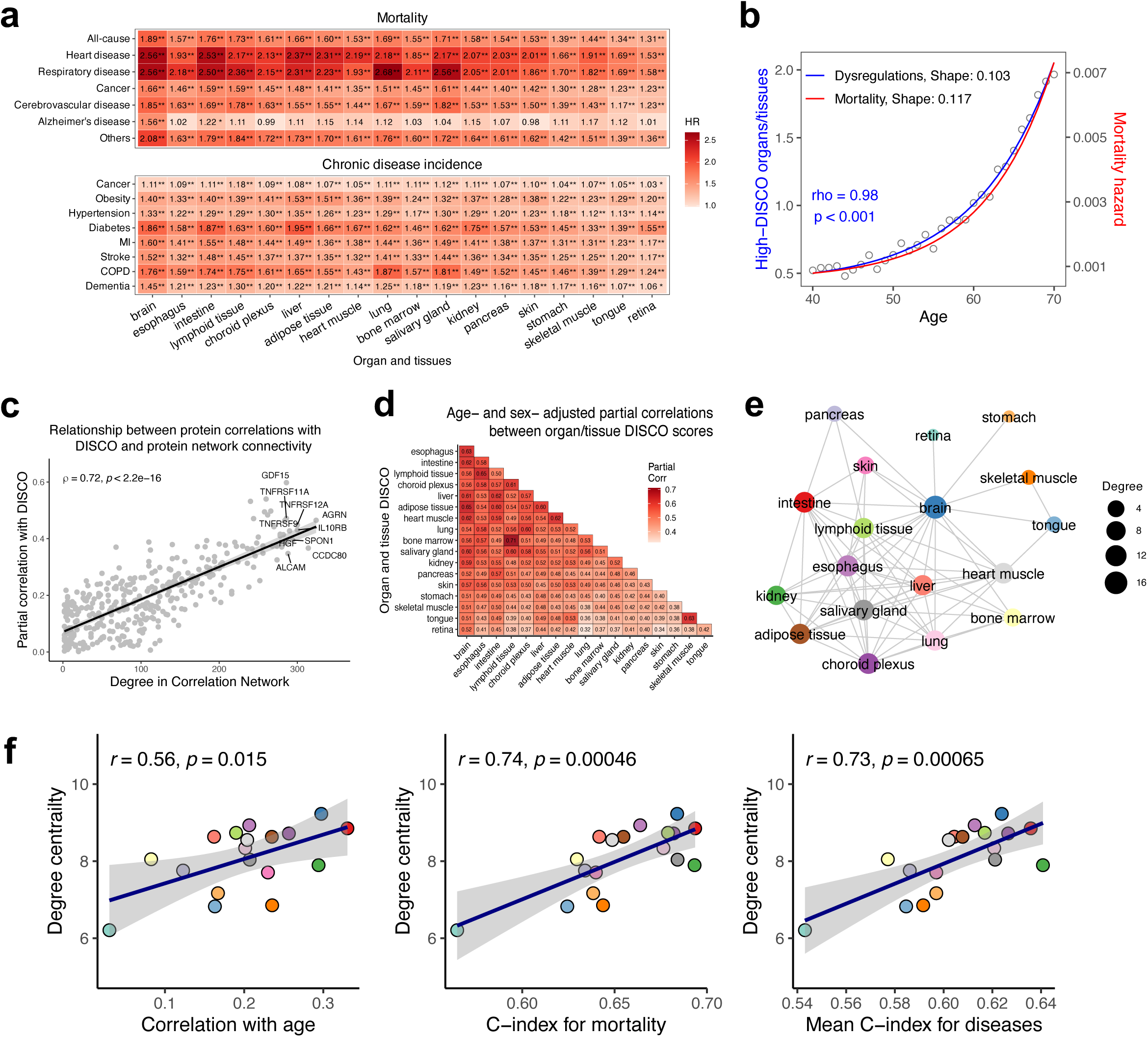
Multi-organ entropy profiling by DISCO. Heatmaps (**a**) present age- and sex-adjusted hazard ratios (HRs) for all-cause and cause-specific mortality, and chronic diseases (significances: **: p<0.01; *: p<0.05). (**b**) The ratio of observed/expected high-risk (top 10% DISCO scores) organs and tissues (black circles) increases exponentially with age; the blue line shows a fitted Gompertz-Makeham model. For comparison, the red line shows a Gompertz-Makeham fit to mortality. A scatter plot (**c**) displays biomarker network degree versus DISCO partial correlations (x-axis: network degree; y-axis: partial correlation with DISCO, adjusted for age and sex). A heatmap (**d**) shows the partial correlations between DISCO values of 18 organs/tissues, adjusted for age and sex. A K-neighbor (K=3) network (**e**) visualizes strong inter-organ/tissue entropy links among organs/tissues. (**f**) Scatter plots show relationships between organ/tissue degree centrality (sum of correlations with all other nodes) and three key entropy metrics, with node colors matching panel D. Significance: correlation coefficients and p-values (Pearson) are shown.

We then asked whether the proteins or organ/tissue DISCOs that were most central in networks played more important roles in aging. At the protein level, there was a strong correlation between degree in correlation network and partial correlation with DISCO (ρ = 0.72, p < 0.0001; **Fig. 4c**). Indeed, the single protein with the strongest partial correlation with DISCO was GDF15 (ρ = 0.60, p < 0.0001), which consistently emerges as the top hit in proteomics studies of health and aging^20–22^. GDF15 is thought to signal energetic stress that accumulates during aging and thus to be a broad marker of accumulated dysregulation^23,24^; its association here validates this interpretation of DISCO. Next, we generated a correlation matrix of organ/tissue DISCO scores. Age- and sex-adjusted correlations ranged from 0.32 to 0.71, mean = 0.49, showing much stronger associations than organ protein aging scores in recent papers^8,9^. Consistent with the broad-spectrum prediction of health outcomes (**Fig. 4a**), this suggests that organ/tissue DISCO scores, despite having no overlap in component proteins, are much more convergent across organs/tissues than organ aging scores. We then leveraged this correlation matrix to generate a network of organ/tissue DISCO scores (**Fig. 4d**). Degree centrality based on this network (**Fig. 4e**) was strongly associated with age correlations and C-indices for mortality and disease incidence (**Fig. 4f**). In other words, the organ/tissue DISCO scores that most closely correlate with other DISCO scores are also those that best capture aging, disease risk, and mortality risk broadly. These results were validated through sensitivity analysis across protein selections and model parameters (Extended Data Fig. 9-10).

These findings suggest that additive metrics of aging such as organ-specific -omics clocks and integrative metrics of aging such as DISCO provide profoundly different insights into the nature of the aging process. While organ aging clocks do show that there are clear, organ-specific directional changes with age that are relevant for health and disease^8–10^, DISCO identifies a systemic layer of entropy that is shared across organs and tissues. In this shared layer, dysregulatory dynamics within a given tissue can reflect the overall state of the organism—strongly predicting mortality from diverse, seemingly unrelated causes and providing entropy information that is relatively independent of chronological age (**Fig. 3a**, Extended Data Fig. 3).

We had predicted that DISCO-based cross-organ predictions of health outcomes would be significant but weak, but this was not the case. For example, taking into account the scale of DISCO, the hazard ratios of 2.56 for heart or respiratory disease mortality per unit brain DISCO translate into ∼43-fold higher risk for someone in the top 2.5% of DISCO compared to someone in the bottom 2.5%. Differences in scale make direct comparisons with Oh et al.’s organ age gaps difficult, but this is expected to be roughly on par with the HR=2.37 effect of a heart organ age gap on congestive heart failure, and substantially stronger than the impact of any other organ age gap on congestive heart failure (most are not significant)^8^.

Our results also establish entropy broadly and DISCO specifically as a promising all-purpose metric of aging and health. It provides strong prediction of mortality, frailty, and incidence of every chronic condition tested. For example, for mortality prediction, DISCO combined with age slightly underperforms the best epigenetic clock (GrimAge; C-index = 0.751 vs. 0.759, within the margin of error), despite the fact that GrimAge was explicitly trained as a mortality predictor^18^, whereas DISCO represents an unsupervised approach solely based on perturbations to the correlation structure of the markers. Compared to DM^5,25^, DISCO either performs equivalently (when applied to clinical biomarkers to predict mortality) or substantially better (when applied to -omics data and/or to predict incidence of chronic conditions).

We have developed an R package to facilitate calculation of DISCO in any dataset with appropriate data. Like DM^26^, one of the challenges is that DISCO cannot be calculated on a single sample without the use of a reference population in which to calculate the correlation matrix. We publish and leverage the relevant correlation matrices for both clinical biomarker and -omics data in the datasets used here, thereby allowing calculation of DISCO with this package for anyone with access to those biomarkers or a subset. While this is appropriate for use in a general population, caution should be exercised in applying it to unusual patient subsets. For example, our general reference populations would not be appropriate for use in understanding kidney disease progression in diabetics versus non-diabetics. Interestingly, while we have used organ and tissue DISCO scores here to demonstrate the cross-system integration of entropy, our findings suggest that such specific scores may not have much value in targeted applications, and that a single overall DISCO metric may be sufficient in most cases.

Our findings have several important implications. First, comprehensive measurement of aging^13,14^ should not rely solely on additive or linear aggregations of biological data, but should also incorporate information on the interrelationships of the components and/or the dynamics. The results clearly demonstrate that entropy measured by DISCO predicts disease and mortality more effectively than existing aging metrics, suggesting it may more closely reflect the fundamental nature of aging. From a whole-system perspective, DISCO captures the organism’s integrated dynamical degradation during aging, thereby offering a novel angle for system-level approaches to slow aging and improve health^27^, as interventions that fail to slow or reverse entropy and dysregulation may not generate meaningful or lasting improvements in health^28^. Second, interventions to improve health should be designed so as to maintain or restore these interconnected homeostatic mechanisms^29^. For example, physical activity is broadly understood to trigger multiple systemic changes that bring biological and physiological systems into general alignment for health^30^; such an approach is likely to be more promising than multiple piece-meal approaches that target specific molecules and pathways without attention to how they integrate.

Overall, we have demonstrated that signatures of entropy with aging are strongly linked to mortality and other health outcomes and are widely concordant across organs and tissues, strongly predicting even mortality and disease originating in other organs and tissues. Future work should attempt to integrate this with network measures of aging^31,32^ and expand DISCO into other data modalities. For example, a recent application of DM to ECG data showed a strong predictive signal for fracture risk^6^; DISCO could equally be tested in such data types and might eventually lead to signals of entropy across biological scales. An important open question is whether DISCO applied to such diverse data types would show the same level of interconnections demonstrated here. Future work should also identify the sequence and drivers of entropy accumulation, and whether aging is initiated by such accumulation alongside a decline in repair mechanisms and resilience.

## Methods

### Study populations

Our study used the data of NHANES 1999-2018, UKB, CHARLS, CLHLS, and RuLAS. The US NHANES is a nationally representative cross-sectional survey of civilian living in the US, approved by the National Center for Health Statistics (NCHS) Ethics Review Board^33^. The UK Biobank is large-scale prospective cohort that collected data from over 500,000 participants across 22 centers in England, Scotland, and Wales. UKB received ethics approval from the North West Multicenter Research Ethics Commitee^34^. The CHARLS is an ongoing prospective population-based longitudinal cohort study of middle-aged and older Chinese adults. CHARLS was approved by the Ethics Review Board of Peking University, which was conducted in accordance with the Declaration of Helsinki and other relevant guidelines and regulations^35^. The CLHLS is a nationwide longitudinal study of old-aged Chinese population. The project was approved by the Biomedical Ethics Committee of Peking University, China (IRB00001052-13074)^36^. The Rugao Longevity and Ageing Study (RuLAS) is a population-based prospective study, which consisted of a longevity cohort and an aging cohort in Rugao, China^37^. The RuLAS was approved by the Human Ethics Committee of Fudan University School of Life Sciences. All participants provided written informed consent. This study followed the Strengthening the Reporting of Observational Studies in Epidemiology (STROBE) reporting guidelines for cohort studies.

### Collection of clinical biomarkers

Here, we use as many clinical biomarkers as possible for each cohort, based on what is accessible. In NHANES, 24 biomarkers of blood biochemical assays (glucose, albumin, etc.) and complete blood count tests (NEU%, LYM%, etc.) were collected. In UKB, 58 biomarkers of hematological assays and biochemistry tests measured in the blood sample were collected in this study. In RuLAS, fasting blood samples were collected for laboratory tests (physical, hematological, and biochemical biomarkers). In RuLAS, 57 biomarkers of Wave 2 (2016) and 63 biomarkers of Wave 4 (2019) were enrolled for analysis, respectively. For CHARLS, both the Wave 1 (2011) and Wave 3 (2015) collected blood samples for complete blood count (CBC) tests and blood-based bioassays, in which 16 biomarkers were measured for the participants. For CLHLS (2014 wave), 30 biomarkers of CBC, blood biochemical examination and urine routine test were involved in DISCO analysis. The details of biomarkers were summarized in **Supplementary Tables 1-6**.

### Collection of proteomics biomarkers

In UKB (2006-2010), the raw plasma proteomics data (Olink platform) contained 2923 proteins from 53,014 participants. In quality control, we removed proteins with more than 10% missing data and participants with more than 50% missing data, resulting in 1461 protein biomarkers on 47,859 participants for analysis. To reduce the computational cost and remove unconcerned variables, we used LASSO-cox regression model (five-fold cross validation) to extract proteins related to mortality risk. We selected the penalty as the model reached the highest C-index for survival analysis. As a result, 367 proteins were obtained through five-fold cross-validation. For the union analysis, 47,173 participants with 58 clinical biomarkers and 367 proteins were enrolled. After feature selection, 396 features (39 biomarkers and 357 proteins) were retained (**Supplementary Tables 7-8**).

### Organ-specific proteomics analysis

The organ and tissue enriched and enhanced proteins were obtained from the Human Protein Altas (https://www.proteinatlas.org). Consequently, proteins from the proteomics analysis were assigned as appropriate to one of 36 tissues. We selected the organs/tissues with more than 40 such proteins for DISCO analysis, and sex-related organs were removed (placenta, ovary, testis). Following the feature selection in previous studies^8,9^, the organ/tissue specific and enhanced proteins were pruned using Lasso-cox regression models with the penalty as the model reached highest C-index (**Supplementary Table 9**). To define organ-specific high-risk DISCO, we employed quantile-based thresholds to estimate parameters of the Gompertz-Makeham (GM) model, which captures age-dependent increases in dysregulation risk. GM parameters stabilized at thresholds ≥10% (Extended Data Fig. 9a), establishing this as the critical cutoff. We then computed an adjusted “number at risk” by dividing the observed age- and individual-averaged number of high-risk DISCO organs by the expected value under uniform distribution assumptions. The expected value was defined as the total number of organs multiplied by the 10% threshold. For pairwise correlation analysis of inter-organ DISCO levels, we first removed overlapping proteins to eliminate redundancy and recalculated DISCO values along with their Pearson correlation coefficients. This preprocessing step ensured that the analysis focused on unique contributors to homeostatic dysregulation across organs. In addition, we performed a comprehensive sensitivity analysis through the following protocol: (1) Protein Sampling. First, organs/tissues were ranked in ascending order based on their protein counts. Following this ranking, 40 proteins were randomly selected from each organ/tissue, a process repeated across 200 independent iterations across 200 independent replicates; (2) Non-overlap constraint. Proteins were sampled under a strict exclusion rule to prevent inter-sample overlap, ensuring independence of observations. DISCO values were computed for each replicate and each organ/tissue. Finally, we performed global correlation analysis on the mean DISCO profiles to identify significant correlations between organs/tissues.

### Collection of metabolomics and microbiota biomarkers

In RuLAS Wave 4 (2019), we collected blood serum metabolomics (profiling through nuclear magnetic resonance) and gut microbiome data^38^. For metabolomics data, 313 metabolites and related variables (amino acid, low-density and high-density lipoproteins, derived ratios, etc) from 2120 participants were collected. Due to strong correlation and collinearity of the metabolomics data, we used principal components analysis to reduce the redundancy and extract the primary information. The first 50 principal components explaining over 95% of the variance of the original variables, were used for DISCO analysis (**Supplementary Table 10**). For gut microbiome data, we selected the microbial species with missing abundancy less than 95% across all participants. Next, 251 microbial species of 1525 participants were used for analysis. We used the metabolomics data to measure the metabolic homeostatic dysregulation, and 27 microbial species correlated with metabolic homeostasis were selected (Spearman p value < 0.05, **Supplementary Table 11**). Due to the sparse distribution of microbial abundance across individuals, the covariance matrix for the microbiome data was computed using Spearman correlation. DM analyses followed the same analytical workflow as DISCO, including biomarker selection, dimensionality reduction, and statistical significance. A total of 30 microbiotas correlated with metabolic homeostasis (defined by DM) were selected, which were highly consistent with those of DISCO with 26 microbiotas overlapped. DM analyses adhered to the same analytical workflow as DISCO, encompassing dimensionality reduction, biomarker selection, and statistical validation. A total of 30 microbiota associated with metabolic homeostasis (as defined by DM) were identified, with 26 overlapping taxa (86.7% consistency) shared between DM and DISCO.

### The calculation of DISCO

The Euclidean Distance of Covariance (DISCO) was defined as the squared sum of differences between the elements of the perturbed covariance matrix and the reference covariance matrix. Assuming that there were *m* biomarkers, DISCO was derived using Pearson correlation coefficients (PCCs) from two groups: a reference group with n samples (denoted as *ref*_*n*_) and a perturbed group with n+1 samples (denoted as *ref*_*n*+1_). Then, we added a weighted score to the component elements of the DISCO, aiming to enhance the weight of aging biomarkers. The DISCO metric is calculated as:

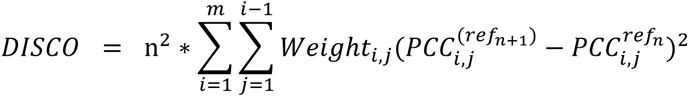

and 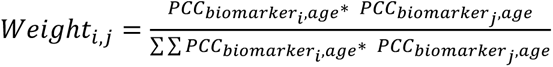

The 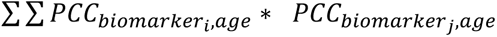 was the absolute value of quadratic sum of PCC between biomarker i, j, and age, working as a scale factor. The raw distribution of DISCO was right-skewed, and we used log transformed DISCO in downstream analysis. The relation of DISCO to entropy was analyzed in the **Supplementary materials**.

### Simulation analysis

To evaluate DISCO as a measure of entropy, we conducted comprehensive simulations comparing it against Mahalanobis distance (d_M_), entropy and KL distance (d_E_). Traditional entropy metrics like Shannon entropy face fundamental limitations in biological applications: they require discrete data distributions, measure population-level rather than individual-level entropy, and cannot capture multivariate correlation shifts critical to aging processes. We therefore developed three approaches quantifying individual entropy as multivariate deviation from a healthy reference distribution: d_M_ (assuming Gaussian distribution), d_E_ and d_KL_ (conceptually appealing but functionally equivalent to d_M_ under Gaussian assumptions), and DISCO (distribution-free and specifically designed to detect correlation structure changes). Our simulation framework employed multivariate Gaussian distributions with Toeplitz covariance matrices (ρ=0.1-0.7) to model systems with dimensionally decaying correlations. We generated 2D and 10D datasets (n=1,000) and evaluated distance metrics across systematic grids in the original and principal component spaces. All metrics were normalized to [0,1] for cross-comparison. Survival simulations modeling mortality risk (Cox, Exponential, Gompertz, and Weibull distributions) with quadratic hazard functions, DISCO outperformed alternatives under strong correlation structures regardless of variable dimensionality (10, 50, 100) or noise level (σ=0.2, 1). Critically, DISCO showed superior sensitivity to fine-scale differences at low entropy levels, suggesting particular utility for detecting early-stage dysregulation in aging processes where correlation structures remain largely intact but begin showing initial signs of disintegration.

### Assessment of clinical outcomes

In NHANES, death information was based on linked data from records taken from the National Death Index (NDI) through December 31, 2019, provided through the Centers for Disease Control and Prevention. In UKB, death information was obtained through death certificates held within the National Health Service (NHS) Information Centre (England and Wales) and the NHS Central Register (Scotland) to November 30, 2022. We used the International Statistical Classification of Diseases, 10th, to define causes of death. The cause-specific mortality included mortality of malignant neoplasm, heart disease, cerebrovascular disease, respiratory disease, Alzheimer disease, diabetes, and others. In addition, diagnosed dates of incident chronic disease in UKB were also collected, including obesity, hypertension, diabetes, cancer, myocardial infraction, heart failure, stroke, chronic obstructive pulmonary disease (COPD), and dementia. In RuLAS, dates of death for the participants were obtained from the Bureau of Civil Affairs of Rugao. For RuLAS Wave 2 (2016) and Wave 4 (2019), 4-year survival and 2-year survival data were calculated, respectively. In CHARLS, records on death were obtained at the biennial follow-up. Because the exact date of death was not available, the point of the last follow-up visits was defined as the time of death. In CHARLS, we used frailty index (FI) to evaluate the frailty status using chronic diseases and functional limitations were collected for frailty assessment (**Supplementary Table 12**). Participants were divided into robust (FI<0.1), prefrail (0.1<=FI<0.25) and frail groups (FI>=0.25). In CLHLS, death information was collected from the official death certificates when available. Otherwise, the information was obtained from next-of-kin and local residential committees.

### Benchmark and robustness analysis

DISCO’s predictive performance for mortality and disease outcomes was benchmarked against DM. We further analyzed DNA methylation aging clock data (**Supplementary Table 13**) from the NHANES 1999–2002 cohort, which initially included 4,449 participants. After excluding 1,917 individuals with incomplete methylation data, 2,532 participants were retained for analysis. Over a median follow-up of 17.2 years (IQR: 9.3–18.8), 1,361 all-cause mortality events were recorded. All-cause mortality served as the outcome for comparing DISCO and DM against established aging clocks. To assess DISCO’s robustness, we evaluated its distribution stability and feature correlation under varying reference group sizes and biomarker selection strategies, conducting 50 randomized replicates for each parameter configuration. We further examined its performance across different age-stratified reference group definitions and alternative matrix distance metrics. Additionally, DISCO’s predictive capacity for mortality was systematically tested with incremental biomarker inclusion, with mean performance metrics calculated over 50 randomized replicates at each step. Finally, leave-one-biomarker-out analysis validated its robustness to individual feature selection, with performance distributions estimated via 50 bootstrap iterations.

### Statistical analysis

Participants were classified into quartiles through DISCO/DM, with the first and fourth quartiles representing individuals at lowest and highest risks, respectively. Kaplan-Meier survival curves were then plotted to compare the predicted survival probabilities. We utilized Cox proportional hazard models to estimate the hazard ratios (HRs) and 95% confidence intervals (CIs) of levels associated with mortality/diseases using two models. Model 1: crude model; Model 2: adjusted for age and sex. The association analysis was shown in **Supplementary Tables 14-16**. From the Cox models, each individual’s linear predictor was then used to calculate the concordance index (C-index), a measure of the model’s discriminative ability. Harrell’s C-index is known to be biased in the presence of right censoring^39^, which is present in our datasets. Although Uno’s C-index^40^ corrects for this bias, it relies on the assumption of non-informative censoring, which may not hold in practice. We tested both indices for a subset of analyses (mortality prediction in NHANES) and found nearly identical results. Accordingly, we used Harrell’s C-index throughout to assess the predictive discrimination in survival analysis. For binary outcomes (x-year moratlity, x varied by outcomes), the Area Under the Curve (AUC) was took as a robust metric to evaluate the prediction ability. Cox proportional hazard models were conducted to assess the associations between different biological aging clocks and the onset of chronic diseases, including chronic obstructive pulmonary disease (COPD), dementia, and others. These Cox models were adjusted for age and sex. All statistical analyses were performed using R version 4.3.3.

## Acknowledgments

The data used in this research were obtained from the NHANES, UKB, CHARLS, RuLAS and CLHLS. We would like to thank the workers, researchers, and participants involved in these cohorts.

## Data sharing statement

The data of RulAS are available through reasonable request from the corresponding author. The data from CLHLS are available at https://opendata.pku.edu.cn/dataset.xhtml. The data from CHARLS are available at https://charls.charlsdata.com/pages/data/111/zh-cn.html. The data from the NHANES are available at www.cdc.gov/nchs/nhis/index.htm, and the data from the UK Biobank are available upon application at www.ukbiobank.ac.uk/register-apply. This research was conducted using UK Biobank Resource under Application Number 103791.

## Author contributions

Concept and design: Meng Hao, Hui Zhang.

Acquisition, analysis, or interpretation of data: Meng Hao, Zixin Hu, Shuai Jiang, Jingyi Wu.

Drafting of the manuscript: Meng Hao, Hui Zhang, Alan A Cohen.

Critical revision of the manuscript for important intellectual content: All authors.

Statistical analysis: Meng Hao, Hui Zhang, Yi Li, Yaqi Huang.

Administrative, technical, or material support: Xiangnan Li, Shuming Wang, Meijia Wang, Yaqi Huang, Jiaofeng Wang, Jie Chen, Zhijun Bao, Li Jin.

Supervision: Xiaofeng Wang, Alan A Cohen.

---

**Extended Data Table 1.** Associations of DISCO with mortality across five cohorts.

| Cohorts | Follow-up years | Events/N | HR (95% CI) | DISCO C-index | DM C-index |
| --- | --- | --- | --- | --- | --- |
| NHANES | 10.1 (3.8-13.1) | 3304/27059 | 1.71 (1.68-1.75) | 0.767 | 0.766 |
| UKB |  |  |  |  |  |
| Clinical | 13.8 (13.0-14.5) | 42770/487517 | 1.61 (1.60-1.62) | 0.670 | 0.655 |
| Proteomics |  | 5117/47859 | 2.41 (2.35-2.47) | 0.740 | 0.697 |
| ClinicalProt |  | 5022/47173 | 2.38 (2.33-2.44) | 0.741 | 0.677 |
| Validations |  |  |  |  |  |
| CHARLS | 8.0 (8.0-8.0) | 1096/11601 | 1.26 (1.21-1.30) | 0.689 | 0.670 |
| CLHLS | 3.3 (2.2-3.5) | 828/2213 | 1.27 (1.21-1.33) | 0.651 | 0.605 |
| RuLAS |  |  |  |  |  |
| wave 2 | 4.0 (4.0-4.0) | 190/1821 | 1.33 (1.21-1.46) | 0.650 | 0.604 |
| wave 4 | 2.0 (2.0-2.0) | 124/2120 | 1.45 (1.30-1.62) | 0.678 | 0.662 |

Follow-up years were shown with median (interquartile). HR (95% CI) was estimated adjusting for age and sex, while C-index was calculated based on crude model (mortality ~ DISCO/DM). HR: hazard ratio; CI: confidence interval; DISCO: distance of covariance; DM: Mahalanobis distance;

**Extended Data Fig. 1.**
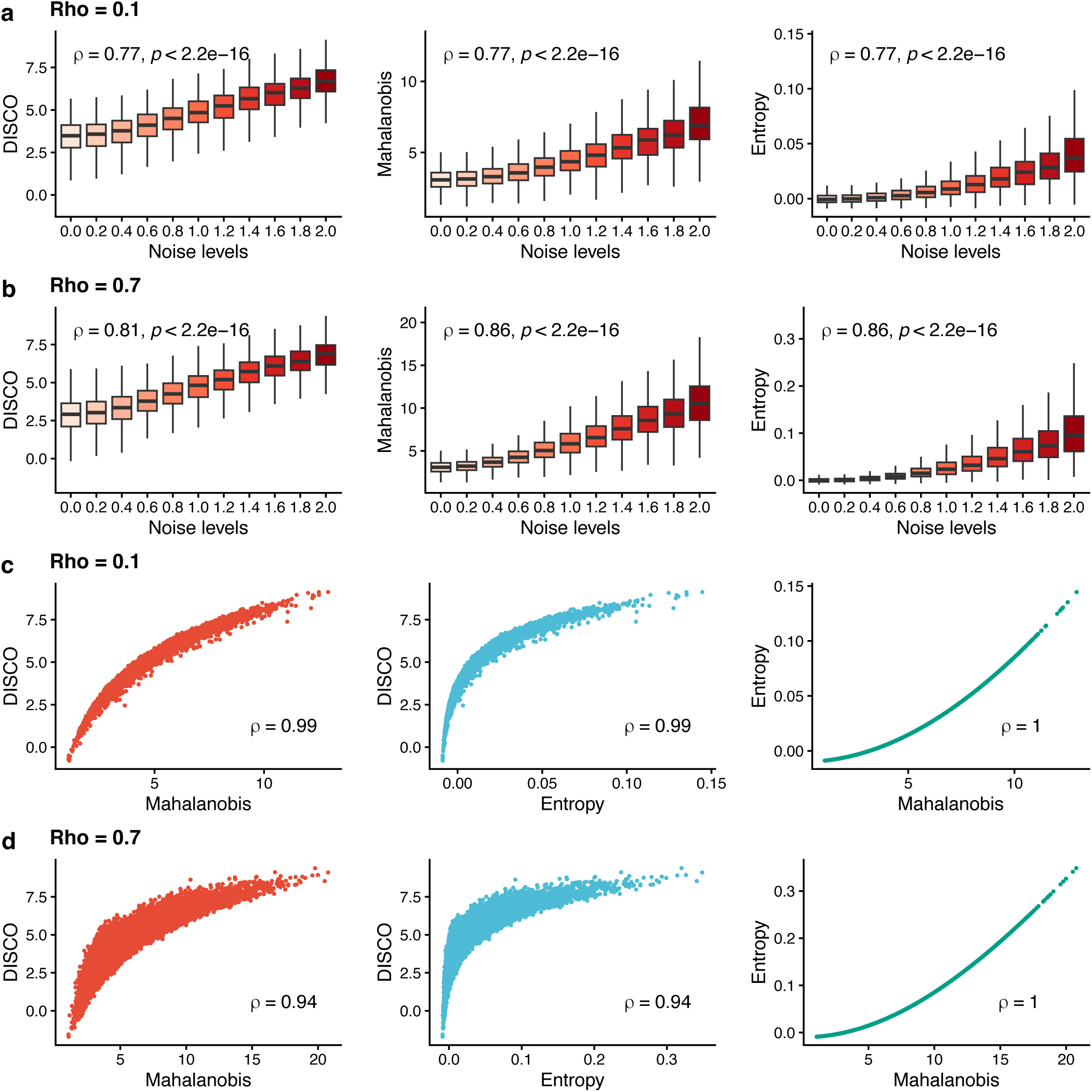
Simulation analysis of DISCO in relation to DM, and Entropy. Boxplots (**a-b**) show increased DISCO with simulated entropy increased systems. Both datasets (10-dimensional) and noises were generated from Gaussian distributions. Datasets were simulated through multivariate Gaussian distributions with Toeplitz covariance matrices (ρ=0.1, 0.7). Scatter plots (**c-d**) of DISCO with DM and Entropy distance. Spearman’s correlation coefficients (ρ) and p-values were shown. DM: Mahalanobis distance; Entropy distance was measured as the entropy differences of the Gaussian systems before and after the additional individual data.

**Extended Data Fig 2.**
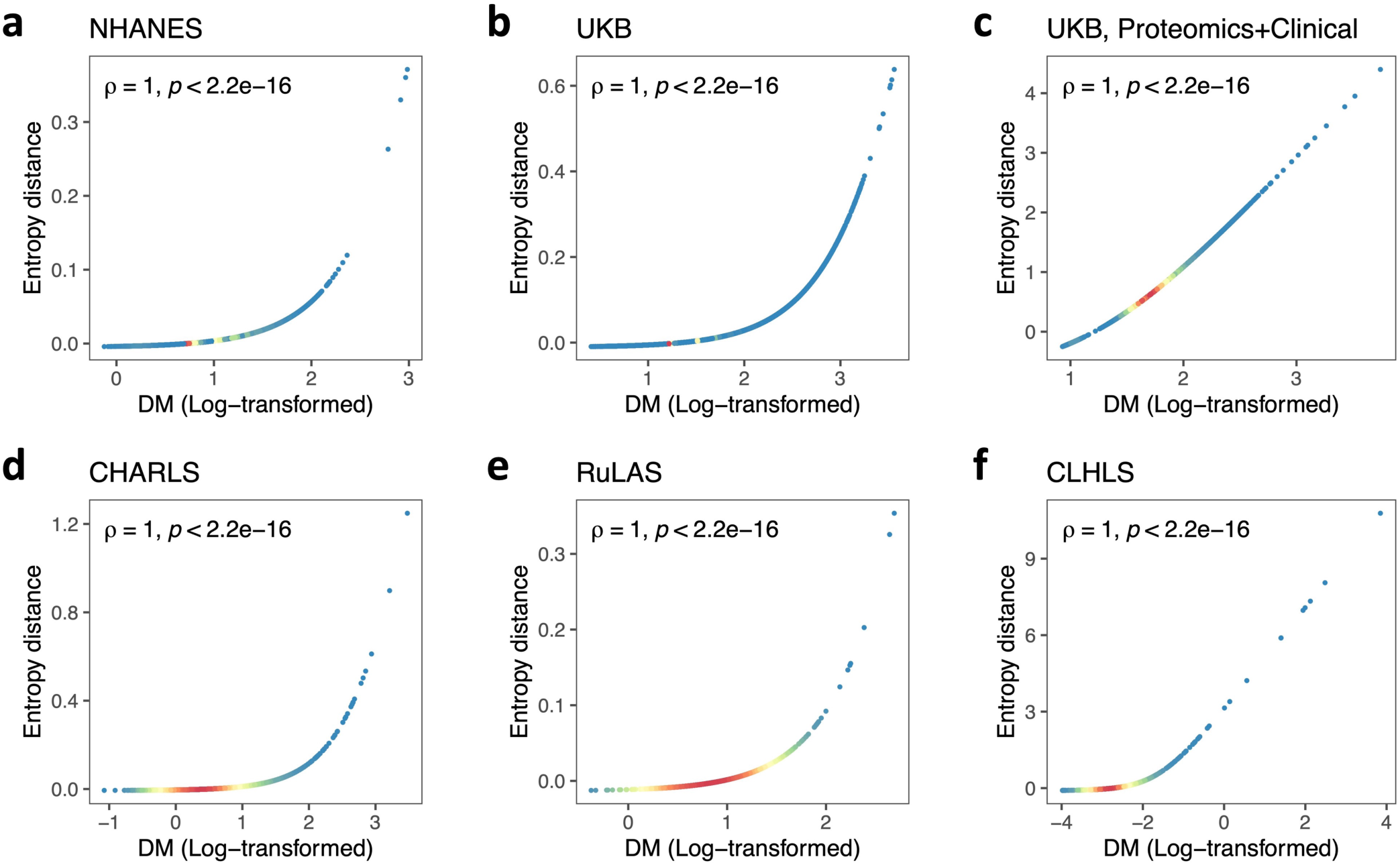
The relationships between Mahalanobis distance (DM) and Entropy distance across real cohort datasets. Entropy distance quantifies the change in population entropy after incorporating an additional individual. Both metrics were derived from the same reference population and biomarker profiles. Panels **a-b**, **d-e**, and **f** are based on clinical biomarkers only, while Panel **c** is based on a combined set of proteomic and clinical biomarkers (425 features total). The red coloration denotes regions with high-density distributions of values, whereas blue signifies areas of low density.

**Extended Data Fig 3.**
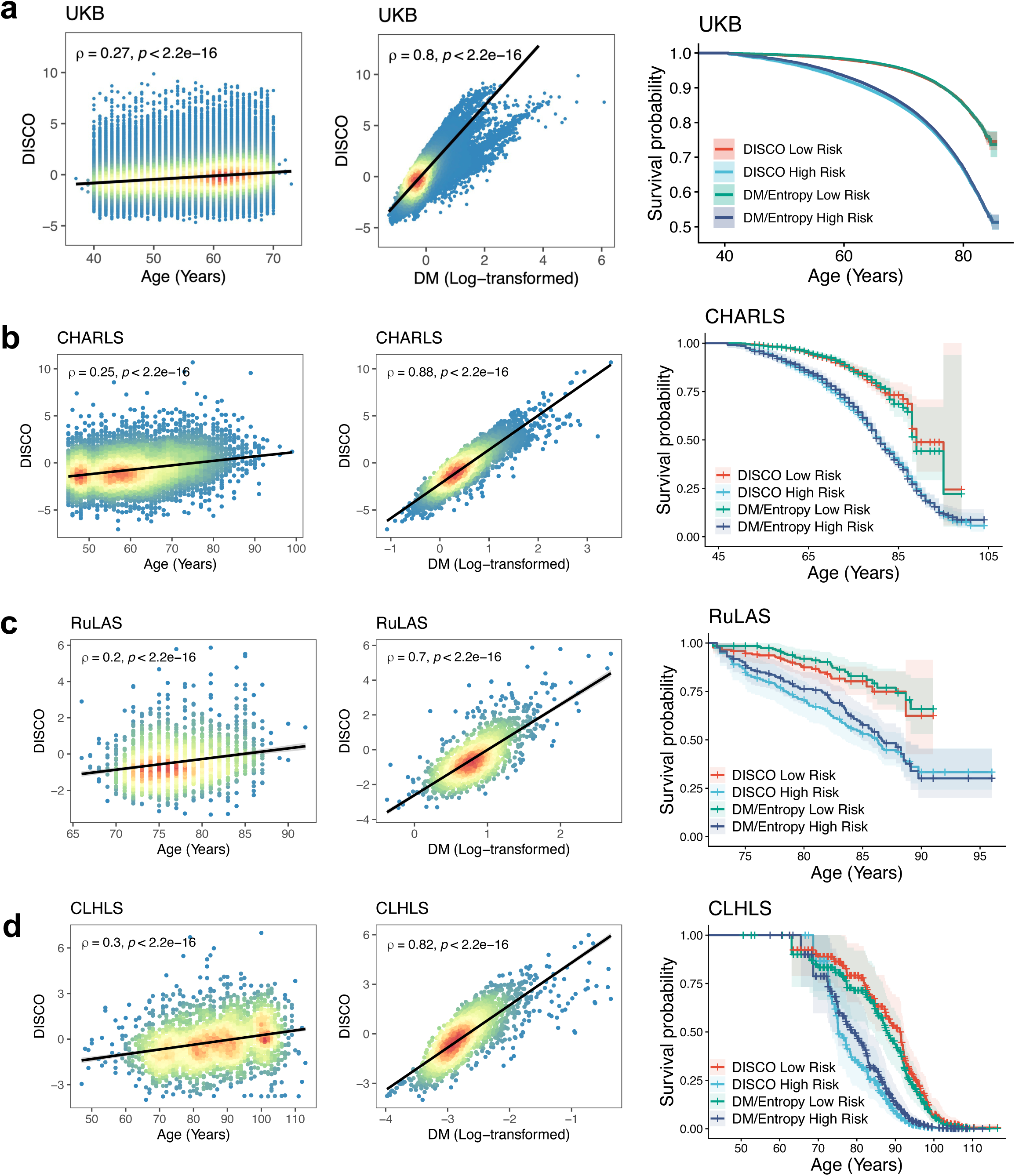
Validations of DISCO in UKB (a), CHARLS (b), RuLAS (c) and CLHLS (d). DISCO versus age (left) and DM (center) across cohorts, with Spearman’s correlation coefficients (ρ) and p-values. Kaplan-Meier curves (right) compared survival probabilities for individuals categorized by low/high risks (first and forth quartiles) according to the DISCO metric and the identical DM/Entropy metric, respectively, using chronological age as the time-scale axis. DM: Mahalanobis distance.

**Extended Data Fig 4.**
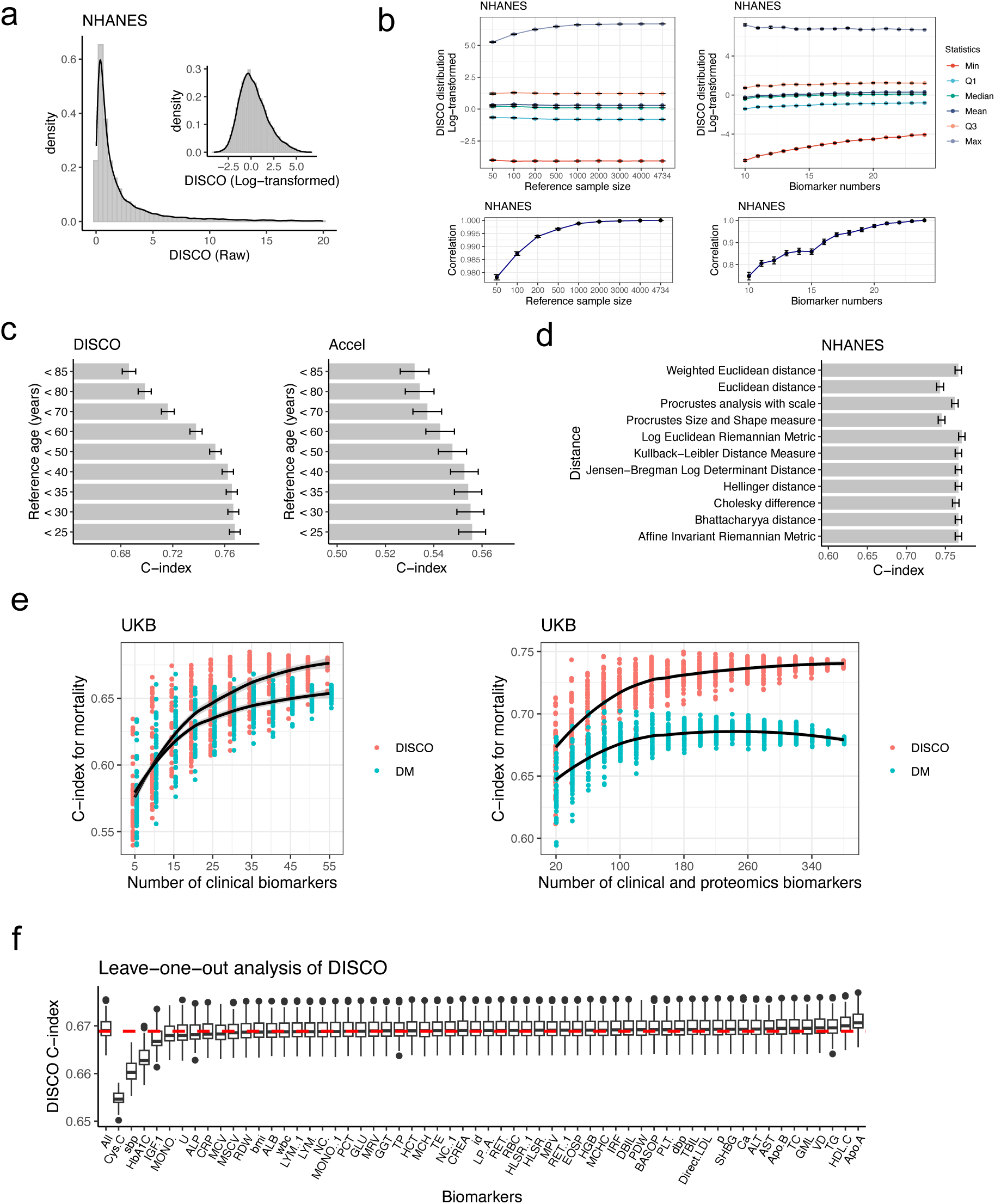
Sensitivity and robustness analysis of DISCO. Density distribution plots (**a**) of raw and log-transformed DISCO values. The DISCO distribution (**b**) across different reference (young adult group) size and biomarker numbers in NHANES. The Spearman correlation of DISCO values for the 50 replicates at each parameter setting were evaluated. Bar plots (**c**) evaluated C-Index performance using reference datasets with different ages in the NHANES. Bar plots (**d**) evaluated C-Index performance using varying distance metrics to calculate DISCO in NHANES. Scatter plots (**e**) show C-Index distribution in predicting mortality through DISCO and DM with randomly selected clinical and proteomics biomarkers in the UKB (50 replicates per biomarker number). Boxplots (**f**) evaluated the stability of DISCO C-index in leave-one-out analysis, in which each biomarker was excluded one-at-a-time (50 replicates per biomarker).

**Extended Data Fig 5.**
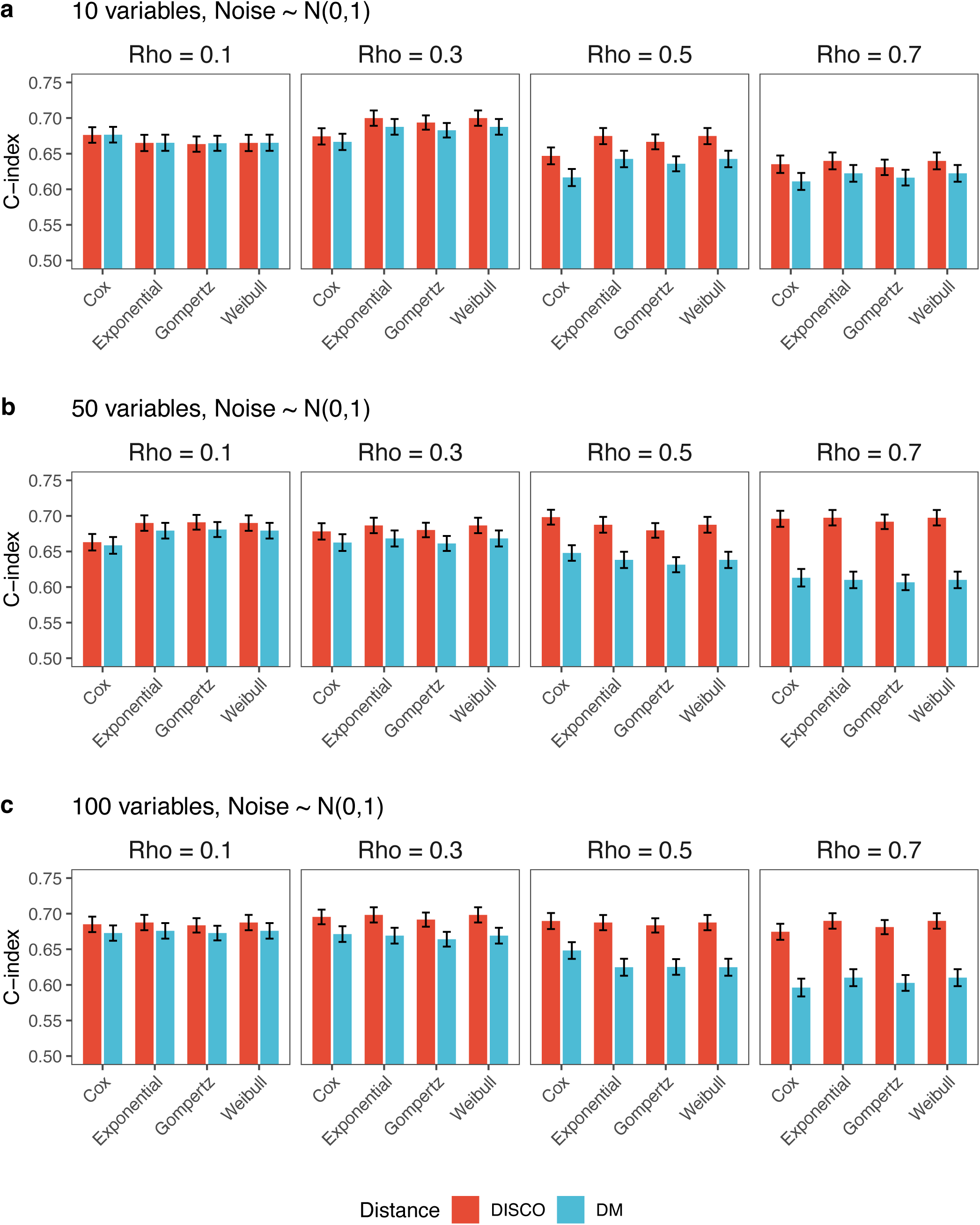
Simulated survival analysis for DISCO/DM. The covariates (n = 10, 50, 100)were generated from Gaussian distributions with Toeplitz matrix Σ (ρ = 0.1, 0.3, 0.5, 0.7). The noise item ɛ was sampled from a normal distribution N(0, σ²), and σ = 1. The survival data was generated under modeling mortality risks (Cox, Exponential, Gompertz, and Weibull distributions).

**Extended Data Fig. 6.**
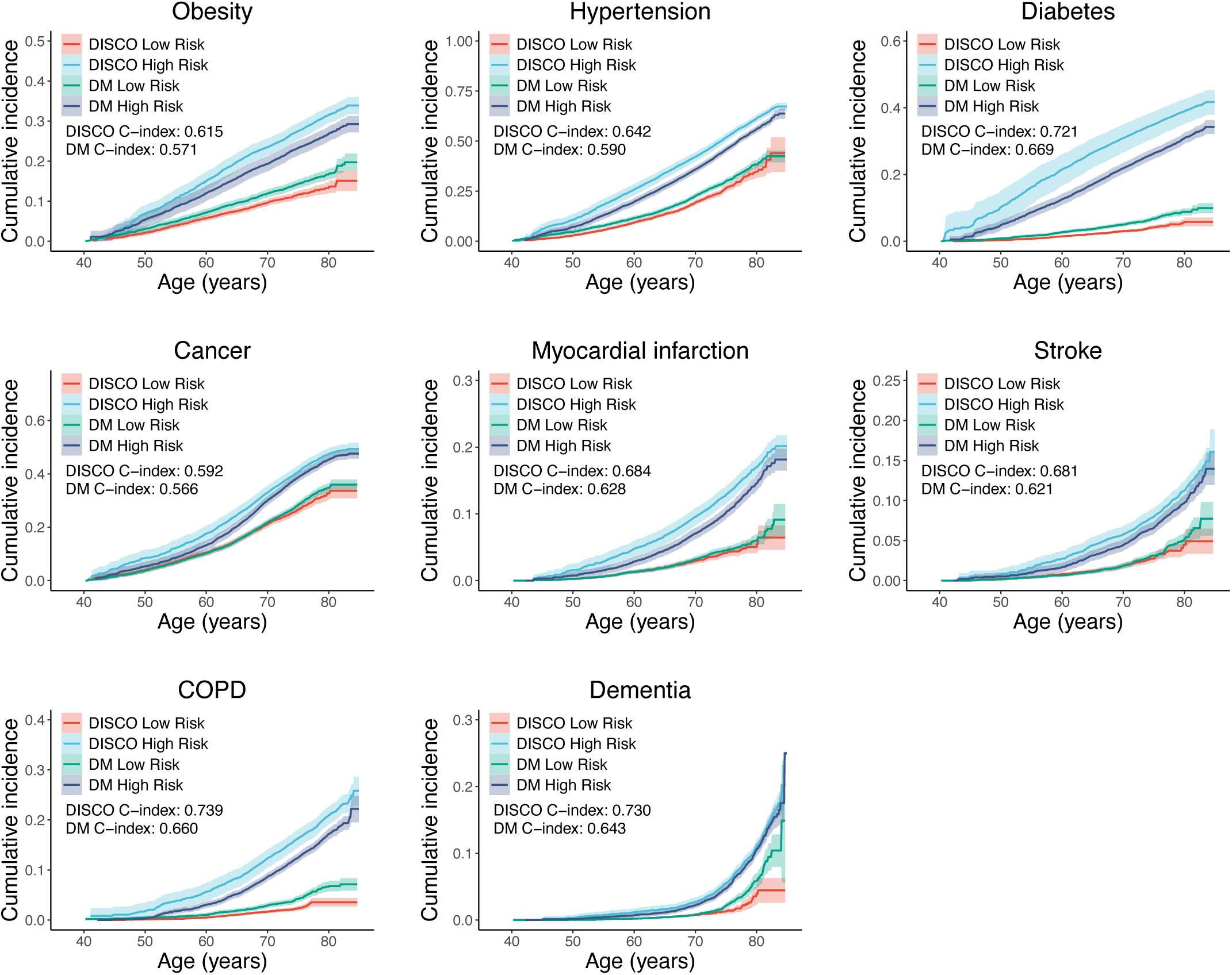
Predictive performance of DISCO for incident chronic disease and frailty. Kaplan-Meier survival curves (**a**) compared the cumulative incidence of chronic diseases (e.g., myocardial infarction [MI], chronic obstructive pulmonary disease [COPD]) in UKB. Individuals were categorized by low (first quartile) and high (fourth quartile) DISCO and Mahalanobis distance (DM) risk groups, with the C-index was shown for each disease.

**Extended Data Fig. 7.**
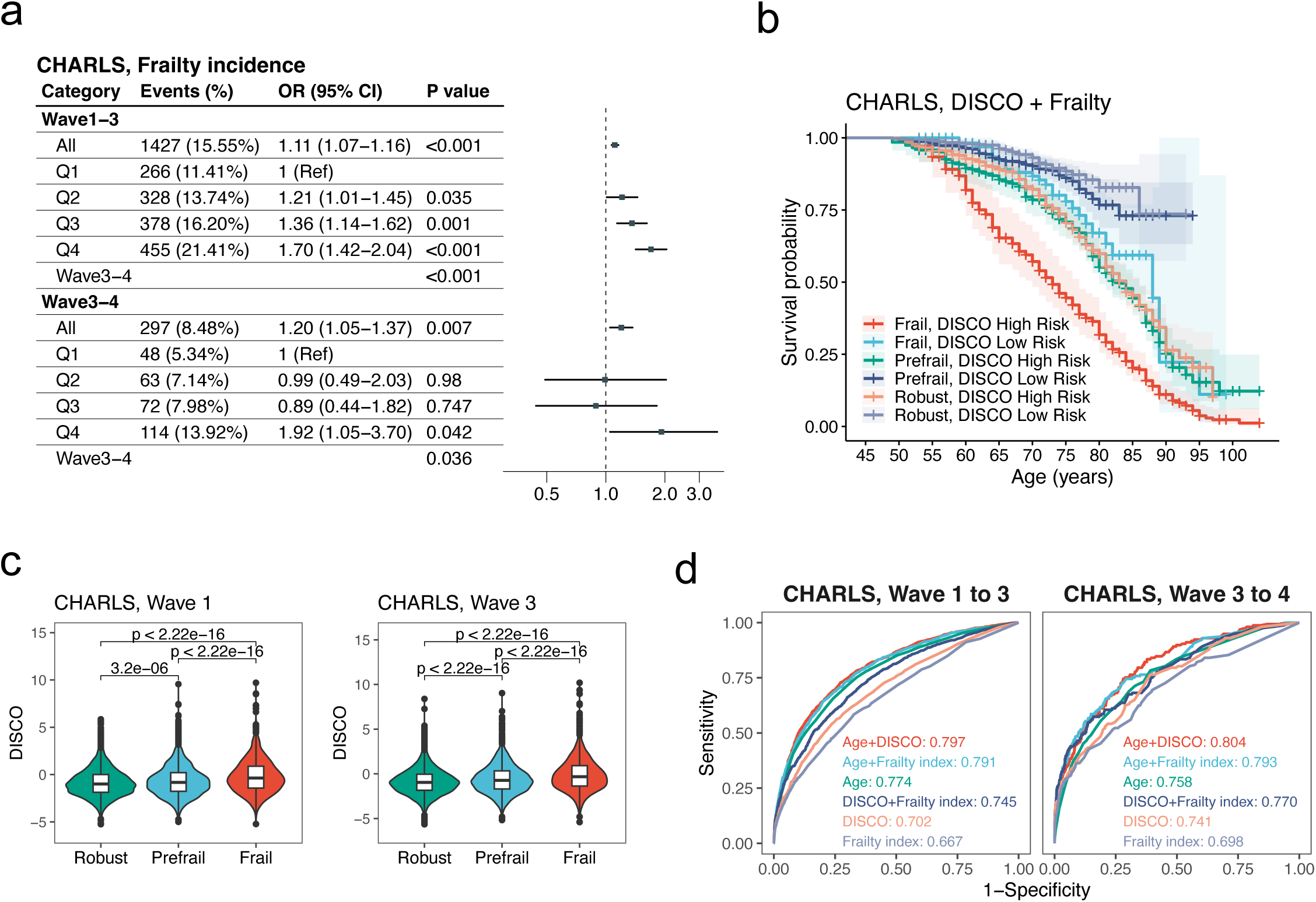
Forest plot (**a**) of odds ratios (ORs, 95% CI) for incident frailty (frailty index ≥0.25, assessed via 32 functional and disease-related items) in the CHARLS cohort, validated across discovery (2011–2015) and validation (2015–2018) waves. Kaplan-Meier survival curves (**b**) stratified participants by DISCO risk quartiles and frailty status. Violin plots (**c**) of DISCO across frailty status which was assessed through frailty index (Robust: frailty index < 0.1; Prefrail: 0.1<= frailty index<0.25; Frail: frailty index >= 0.25). Differences were analyzed using the Wilcoxon tests. Receiver operating characteristic (ROC) curves (**d**) comparing discriminative accuracy of DISCO, frailty index, and age for 4-year (2011 to 2015) and 3-year (2015 to 2018) mortality prediction in CHARLS. The area under the curve (AUC) was shown. ROC: receiver operating characteristic curve; AUC: area under curve.

**Extended Data Fig. 8.**
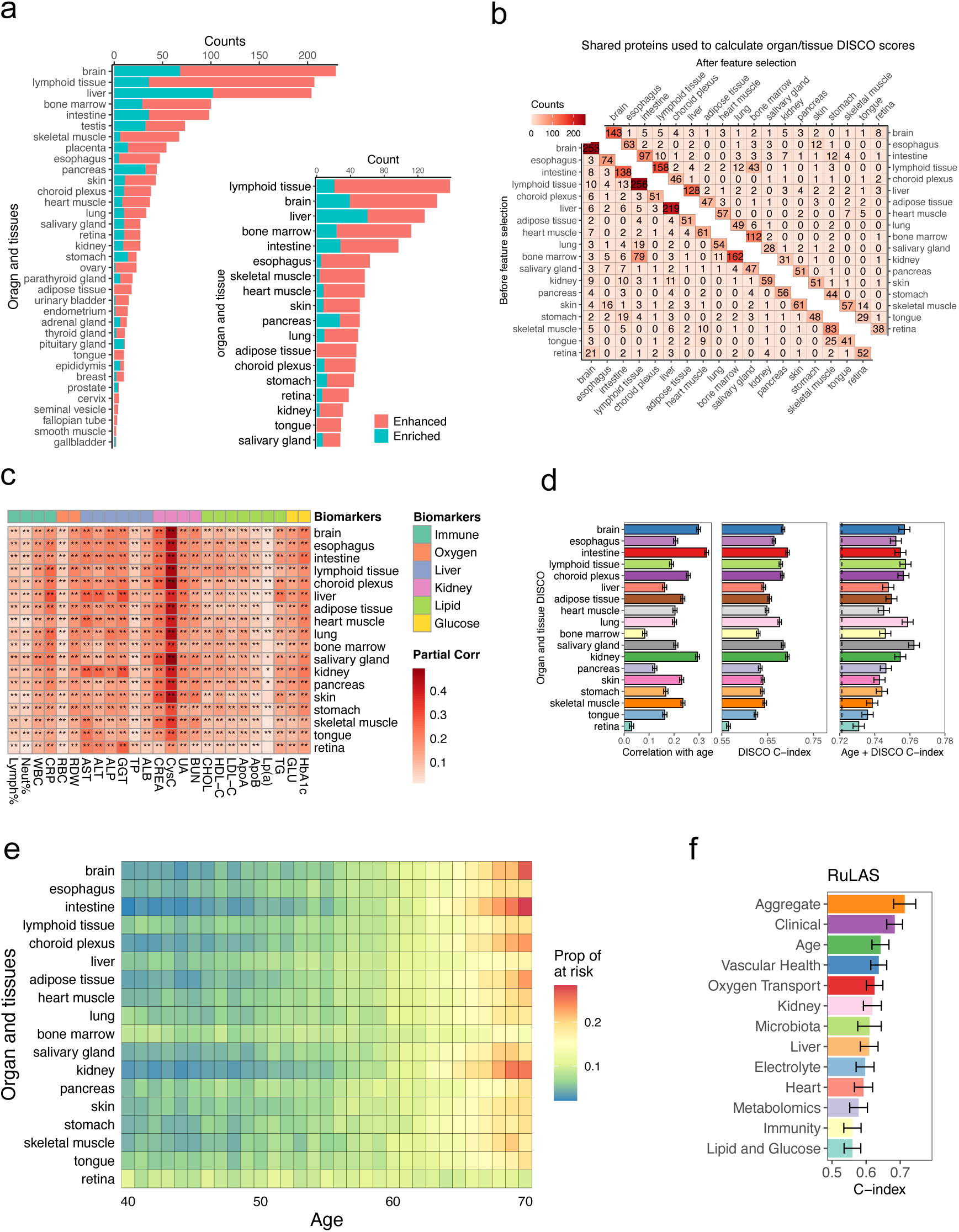
Bar plots (**a**) of organ/tissue-specific protein counts (enhanced: ≥fourfold mRNA in one tissue vs. average; enriched: ≥ fourfold vs. all others) in UKB before (left) and after (right) LASSO selection. Matrix heatmaps (**b**) show the number of shared proteins between pairwise organ/tissue groups before (left lower triangle) and after (right upper triangle) feature selection. A heatmap (**c**) visualizes the partial correlations (r) between DISCO values (18 organs/tissues) and routine clinical biomarkers, adjusting for age and sex. Bar plots (d) show Pearson correlations between organ-specific DISCO and age, C-index of mortality prediction for DISCO alone, and C-index for DISCO + age for 18 organs and tissues. The vertical dashed line indicates C-index of age. A heatmap (**e**) shows age-stratified high-risk organ prevalence (DISCO top 10%). Bar plot (**f**) displays the C-index for 2-year mortality prediction across organ systems defined through clinical, metabolomics and microbiota data in RuLAS. Standard errors (SEs) are shown as error bars in Panel **d** and **f**.

**Extended Data Fig. 9.**
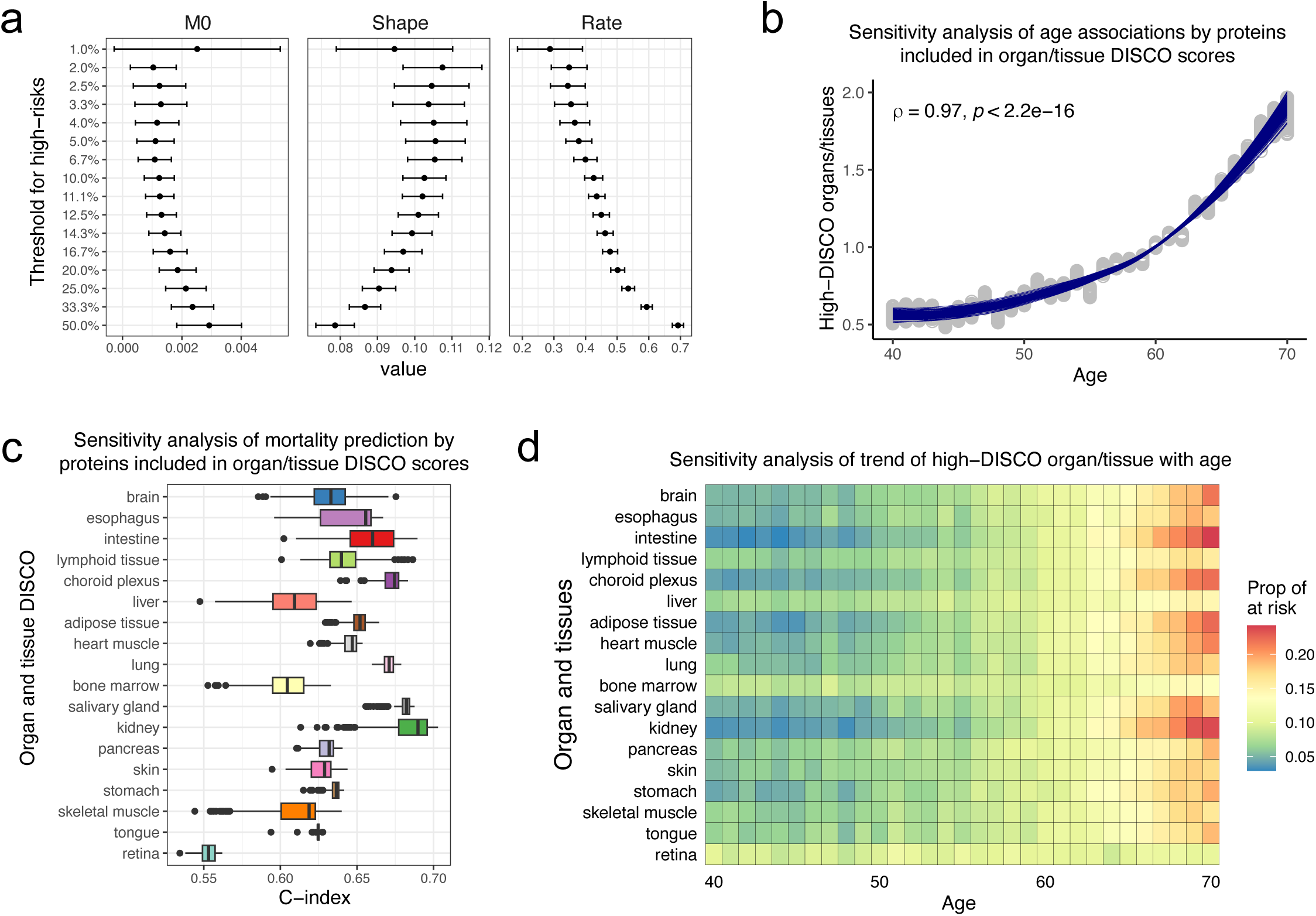
Sensitivity analysis in relation to model parameters and protein selections. (**a**) Parameters of the Gompertz-Makeham model (M_0_, Shape, Rate, y= M_0_ + Rate· *e*^*Shape·Age*^, y is the normalized number of high-risk DISCO organ/tissues) were estimated using a set of quantiles of DISCO to define high-risk entropy across age. Panels **b-d** show the results of sensitivity analysis using randomly selected proteins (40 proteins per organ, 200 replicates). The C-index and multi-organ entropy trends across age were evaluated.

**Extended Data Fig. 10.**
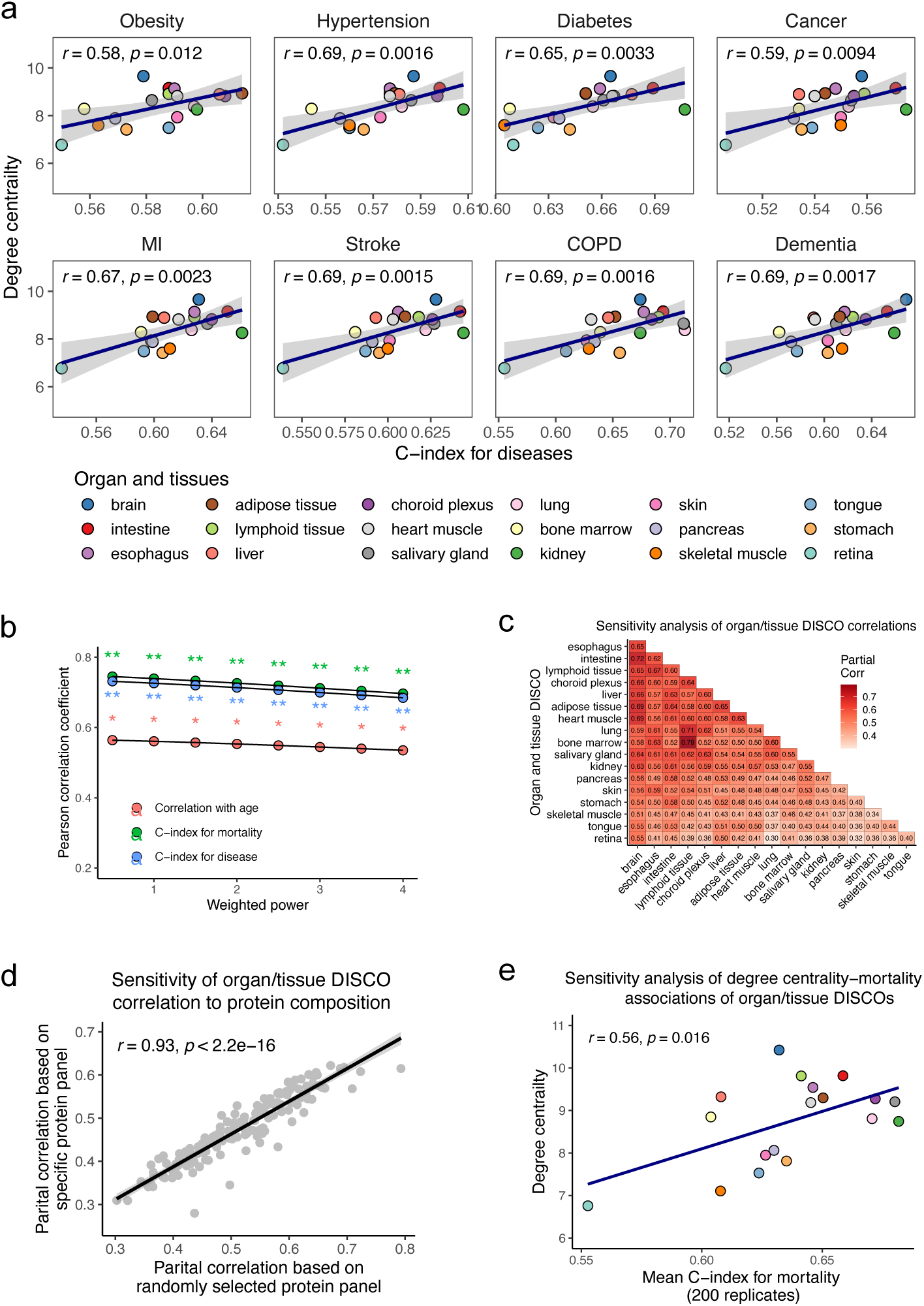
Sensitivity analysis of multi-organ homeostatic dysregulation. Scatter plots (**a**) show degree centrality of organs/tissues versus C-index for predicting chronic disease incidence (Pearson’s r and p values) in UKB. Sensitivity analysis (**b**) of the weighting power parameter used to define degree centrality on the correlations in Fig. 4e. A heatmap (**c**) depicts age-/sex-adjusted partial correlations between organ-specific DISCO values, averaged across 200 replicates of randomly selected protein subsets (40 proteins per organ). The relationships (**d**) of pairwise inter-organ/tissue DISCO partial correlations (y-axis: raw proteomics data adjusted for age/sex; x-axis: 200 random protein subsets). (**e**) Sensitivity analysis (40 proteins per system, 200 replicates) of the association between organ/tissue degree centrality and mortality prediction C-index using DISCO derived from among organ-specific proteins, with node colors matching panel **a**. Statistical significance is indicated by Pearson correlation coefficients (r) and p-values.

