## Supplementary Information for "Human aging reflects increases in entropy across organ networks"

### Supplementary materials of DISCO paper

#### Preface Outline

Entropy is traditionally measured with classical metrics such as Shannon entropy. With ideal data, one could directly measure Shannon entropy in biological data and reate it to various aspects of the aging process. However, several fundamental issues prevent this in most biological data:

(1). Spatial or temporal structure in the data. Alternative formulations of Shannon entropy exist for such situations.

(2). Continuous distributions. Shannon entropy assumes discrete (categorical) data, but many biological parameters vary continuously. Differential entropy is related to Shannon entropy but is not an exact equivalent.

(3). Individual measurement challenge: Shannon entropy and all related methods measure the entropy of a distribution, not a value. However, we are often interested in quantifying the entropy of an individual sample, not a distribution. For example, in typical biomarker data, we might represent individuals as rows and biomarkers as columns; Shannon or differential entropy could be calculated with a single value for each column, but we would actually like to calculate a single value for each row. Accurate measurement is thus infeasible with standard methods for a single individual. Even if one biomarker (e.g., pulse rate) can be sampled precisely, the correlations between it and all other biomarkers cannot, making the complete joint distribution practically unmeasurable.

(4). Limitations of Standard Methods: The standard entropy-related method, e.g, Kullback-Leibler (KL) divergence, requires a reference distribution for comparison. While Shannon entropy cannot conceptually be applied to a single value, it should nonetheless be the case that entropic processes occurring on an individual over time should leave signatures in the individual values of biomarkers or other parameters; such signatures should be increasingly evident and robust in comparison to population distributions and as they become multivariate.

(5). Population-Level Shifts: The "aging-specific" signal is not merely the entropy at a specific age. The mean of the biomarker distribution itself shifts with age, meaning the critical factor is the change in distribution relative to a young, healthy reference population.

**(6). Distances as entropy metrics.** We hypothesize that greater deviations from the reference correspond to higher entropy, implying that distance measures can serve as proxies for entropy. The deviation of an individual from a young, healthy reference population reflects how much the individual alters the overall entropy when included in the population.

Here, we introduced four distance-based methods to quantify entropy as an individual's multivariate deviations from a healthy reference distribution. We evaluated four approaches:

a). Mahalanobis distance ($d_M$): A robust method that effectively functions as a log-likelihood calculation. Its primary limitation is the assumption that the parent distribution is Gaussian.

b). Entropy distance ($d_E$) and KL distance ($d_{KL}$): This method is conceptually appealing as it directly engages with entropy. However, because entropy is difficult to measure directly, it is typically estimated under a Gaussian assumption. Both the $d_E$ and $d_{KL}$ measures quantify the change in entropy when individual data perturbs a reference set; $d_E$ uses a direct entropy difference, while $d_{KL}$ uses relative entropy. In practice, this makes $d_E$ and $d_{KL}$ functionally equivalent to $d_M$, revealing $d_M$'s inherent connection to entropy through individual perturbations. Presenting them as novel "entropy-based" improvement constitutes a straw-man argument, as they offers no practical advantage under the same constraining assumption.

c). DISCO (Distance of Covariance): it does not assume any specific form for the parent distribution. For comparison purposes, its performance can be evaluated under a Gaussian assumption, and we linked it to KL divergence.

**1. Mathematical relationships of the three distances**

The Mahalanobis distance ($d_M$), DISCO ($d_D$), and entropy distance ($d_E$) share a profound mathematical link: all are intrinsically rooted in the covariance matrix (Σ) and correlation matrix (C) of multivariate data. These matrices capture second-order moments, encoding variable relationships and joint dispersion. Their formal definitions reveal deeper connections:

**(1). DISCO**

For a point $x \in R^p$, weights $w_{ij}$ (default $w_{ij} = 1$), augmented dataset $D_{new} = D_{ref} \cup \{x\}$, with covariance $\Sigma_{new}$ and correlation $C_{new}$, correlation matrix $C_{ref} \in R^{p\times p}$:

$$d_D(x) = \log (n^2 * \sum_{i,j} w_{ij} \left( \left[C_{ref}\right]_{ij} - [C_{new}]_{ij} \right)^2 ).$$

DISCO quantifies a shift in correlational structure induced by x.

**(2). Mahalanobis Distance**

For a point $x \in R^p$ and reference data with mean $\mu \in R^p$ and covariance matrix $\Sigma_{ref} \in R^{p\times p}$:

$$d_M(x) = \sqrt{(x-\mu)^T \sum\nolimits_{ref}^{-1} (x-\mu)}.$$

The inverse covariance $\Sigma^{-1}$ transforms this into a covariance-normalized Euclidean distance.

**(3). Entropy Distance**

For a multivariate Gaussian distribution $\mathrm{X} \sim \mathcal{N}(\mu, \Sigma)$, differential entropy is

$$H(x) = -\int_{\mathcal{R}^p} f(\mathrm{x}) \log f(\mathrm{x}) d\mathrm{x},$$

Where $f$ is the probability density function (PDF) of x. For a multivariate normal distribution $\mathrm{X}\sim N(u, \Sigma)$, the PDF is,

$$f(x) = \frac{1}{(2\pi)^{p/2}|\Sigma|^{1/2}} \exp\left[-\frac{1}{2}(x-\mu)^T \Sigma^{-1}(x-\mu)\right],$$

with mean $u \in R^p$, covariance $\Sigma \in R^{p\times p}$, and the $|\Sigma|$ (the determinant of $\Sigma$).

Substituting the Gaussian PDF and simplifying yields

$$H(X) = \frac{p}{2} log\,(2\pi) + \frac{1}{2} log\,|\Sigma| + \frac{1}{2}\int f(x)[(x-\mu)^T \Sigma^{-1}(x-\mu)]dx$$

The expectation term is the trace of the identity matrix：

$$\mathbb{E}[(\mathrm{x}-\mu)^T \Sigma^{-1}(\mathrm{x}-\mu)] = \mathrm{tr}(\Sigma^{-1}\Sigma) = \mathrm{tr}(I_p) = p$$

Thus,

$$H(\mathrm{X}) = \frac{p}{2}\log\,(2\pi) + \frac{1}{2}\log\,|\Sigma| + \frac{p}{2} = \frac{1}{2}[p\log\,(2\pi e) + \log\,|\Sigma|]$$

Similar to DISCO, entropy distance measures the change in entropy when x is added to the reference data. Let $\Sigma_{ref}$ and $\Sigma_{new}$ denote the covariance matrices of the reference and augmented datasets, respectively. The entropy distance is

$$d_E(x) = \frac{1}{2}|\log\det(\Sigma_{new}) - \log\det(\Sigma_{ref})|.$$

This closed-form expression requires X to follow a multivariate Gaussian distribution. For non-Gaussian data, $d_E$ is an approximation.

**(4). KL distance**

We measured the KL divergence as the relative entropy between the new dataset (after adding individual data) and the reference dataset. For two multivariate Gaussian distributions $P\sim N(\mu_p, \Sigma_p)$, and $Q\sim N(\mu_q, \Sigma_q)$, the Kullback-Leibler (KL) divergence is defined as:

$$D_{KL}(P||Q) = \frac{1}{2}\left[tr\left(\Sigma_q^{-1}\Sigma_p\right) + \left(\mu_p - \mu_q\right)^T \Sigma_q^{-1}\left(\mu_p - \mu_q\right) - p + \log\left(\frac{|\Sigma_q|}{|\Sigma_p|}\right)\right].$$

Assume a multivariate Gaussian distribution ($N(\mu_{ref}, \Sigma_{ref})$) of the reference, after adding a new x, it followed $N(\mu_{new}, \Sigma_{new})$. Assume for large reference datasets, adding a new sample x negligibly affects the mean $\mu_{\text{new}} \approx \mu_{\text{ref}}$ and $\mu_{new} - \mu_{ref} \approx 0$. This simplifies KL to

$$d_{KL}(x) = \frac{1}{2}\left[tr\left(\Sigma_{ref}^{-1}\Sigma_{new}\right) - p + \log\left(\frac{|\Sigma_{ref}|}{|\Sigma_{new}|}\right)\right]$$

All four distances derive their properties from Σ or R: Mahalanobi distance uses $\Sigma^{-1}$ to normalize geometric space. DISCO leverages R (standardized Σ) to detect correlation shifts. In essence, Σ acts as the structural backbone, with each distance extracting distinct features: positional deviation (Mahalanobis), correlational patterns (DISCO), and overall scale (entropy, KL divergence).

**2. Eigen-decomposition perspective of the four distances**

Let $\Sigma_{ref} = Q\Lambda Q^T$ be the eigen decomposition of the covariance matrix, where: $Q = [q_1, \dots, q_p]$= eigenvector matrix (orthogonal, ($Q^T Q = I_p$), $\Lambda = \text{diag}(\lambda_1, \dots, \lambda_p)$= eigenvalue matrix ($\lambda_i > 0$)

The three distances can be re-expressed as follows:

(1). Mahalanobis Distance

$$d_M(x) = \sqrt{\sum_{i=1}^{p} \frac{(q_i^T(x-\mu))^2}{\lambda_i}}.$$

It projects the centered point $x - \mu$ onto eigenvectors $q_i$, weights each projection by $1/\lambda_i$ (penalizes directions with small variance). Geometrically, this represents the squared distance after inverse stretching along the principal axes of the distribution.

(2). Entropy Distance

$$d_E(x) = \frac{1}{2}\left|\ln\left(\prod_{i=1}^{p}\lambda_i^{(new)}\right) - \ln\left(\prod_{i=1}^{p}\lambda_i^{(ref)}\right)\right| = \frac{1}{2}\left|\sum_{i=1}^{p} \ln \lambda_i^{(\text{new})} - \sum_{i=1}^{p} \ln \lambda_i^{(\text{ref})}\right|$$

The product $\prod_{i=1}^{p} \lambda_i = \det(\Sigma)$ (the generalized variance) represents the hypervolume of the distribution. It is sensitive to multiplicative changes in eigenvalues, as shown by the equivalent form:

$$d_E(x) \propto \left|\sum_{i=1}^{p} \ln \lambda_i^{(new)} - \sum_{i=1}^{p} \ln \lambda_i^{(ref)}\right|.$$

Geometrically, this measures the log-ratio of the volumes of the confidence ellipsoids.

(3) KL Distance

If the eigenvectors of $\Sigma_{new}$ and $\Sigma_{ref}$ were aligned. Let $\Sigma_{ref} = U\Lambda_{Ref}U^T$ and $\Sigma_{new} = U\Lambda_{new}U^T$.

$$d_{KL}(x) = \frac{1}{2}\sum_{i=1}^{p}\left[\frac{\lambda_i^{(new)}}{\lambda_i^{(new)}} - 1 - log\frac{\lambda_i^{(new)}}{\lambda_i^{(new)}}\right] + \frac{1}{2}\sum_{i=1}^{p}\frac{(u_i^T(\mu_{new}-\mu_{ref}))^2}{\lambda_i^{(new)}},$$

Where $u_i$ represents the i-th eigenvector of the reference covariance matrix ($\Sigma_{ref}$).If the means are approximately equal ($\mu_{\text{new}} \approx \mu_{\text{ref}}$), it simplifies to:

$$d_{KL}(x) = \frac{1}{2}\sum_{i=1}^{p}\left[\frac{\lambda_i^{(new)}}{\lambda_i^{(new)}} - 1 - log\frac{\lambda_i^{(new)}}{\lambda_i^{(new)}}\right]$$

This quantifies purely the change in shape and scale of the confidence ellipsoid, independent of its location. If the covariance structure remains unchanged ($\Sigma_{new} = \Sigma_{ref}$),

$$d_{KL}(x) = \frac{1}{2}\sum_{i=1}^{p}\frac{(u_i^T(\mu_{new} - \mu_{ref}))^2}{\lambda_i^{(new)}} = \frac{1}{2}d_M(x)^2$$

In this special case, the KL distance equals half the squared Mahalanobis distance, establishing a direct mathematical connection between these two measures.

(4). DISCO

Let the reference correlation matrix $C_{\text{ref}}$ have eigen-decomposition $C_{\text{ref}} = U_{\text{ref}}\Phi_{\text{ref}}U_{\text{ref}}^T$,where $U_{\text{ref}} = [\mathbf{u}_1, \dots, \mathbf{u}_p]$ is orthogonal eigenvector matrix, $\Phi_{\text{ref}} = \text{diag}(\phi_1, \dots, \phi_p)$ is eigenvalue matrix. After adding a new sample **x**, the perturbed correlation matrix $C_{\text{new}}$ decomposes as

$$C_{\text{new}} = U_{\text{new}}\,\Phi_{\text{new}}\,U_{\text{new}}^T$$

For the correlation matrix $C_{\text{ref}} = D^{-1}\Sigma_{\text{ref}}D^{-1}$ ,where $D = \text{diag}(\sigma_1, \dots, \sigma_p)$ , the DISCO distance captures the Frobenius norm of the correlation shift

$$d_D(\text{x}) = \log(n^2 \cdot ||\Delta C||_F^2) = \log\left(n^2 \cdot \sum_{i,j}\left([C_{\text{ref}}]_{ij} - [C_{\text{new}}]_{ij}\right)^2\right)$$

Where $\Delta\Phi = \Phi_{\text{ref}} - \Phi_{\text{new}}$, $\Delta U = U_{\text{ref}} - U_{\text{new}}$ It captures correlation shifts arising from two sources: changes in eigenvalues $\Delta\Phi$ and rotations of eigenvectors ($\Delta U$). Geometrically, this corresponds to the Frobenius norm of changes in the eigenstructure of the correlation matrix.

**3. DISCO as a measure of entropy**

As an approximation, DISCO represents a special case of Rényi divergence. When data follow a Gaussian distribution, DISCO distance can be interpreted as a discretized form of Rényi divergence (α=2):

$$D_\alpha(P\|Q) = \frac{1}{\alpha - 1}\log \int p^\alpha(x) q^{1-\alpha}(x)dx$$

Under Gaussian assumptions:

$$d_D(\mathrm{x}) \propto \sum_{i,j} \left([C_{\mathrm{ref}}]_{ij} - [C_{\mathrm{new}}]_{ij}\right)^2 \approx D_2(P_{\mathrm{refl}} \| P_{\mathrm{new}})$$

This formulation emphasizes squared differences, aligning with the sensitivity to second-order moments in Rényi entropy when α=2.

**4.Relationship between DISCO and KL divergence**

The connection between DISCO distance and KL divergence can be established theoretically through perturbation analysis of covariance matrices under Gaussian assumptions. Below is a detailed derivation with key assumptions, and practical applications:

(1). Perturbation analysis

Assume the covariance matrix perturbation induced by a new data point is $\Delta\Sigma = \Sigma_{new} - \Sigma_{ref}$, and $\Delta\Sigma$ is a small perturbation matrix. Taylor expanding the KL divergence:

Trace term Expansion:

For small $\Delta\Sigma$, the inverse of $\Sigma_{new}$ approximates to:

$$\Sigma_{new}^{-1} \approx \Sigma_{ref}^{-1} - \Sigma_{ref}^{-1}\Delta\Sigma\Sigma_{ref}^{-1},$$

$$\text{and } tr\left(\Sigma_{ref}^{-1}\Sigma_{new}\right) \approx p + tr(\Sigma_{ref}^{-1}\Delta\Sigma).$$

Log-determinant approximation:

$$\log|\Sigma_{new}| \approx \log|\Sigma_{ref}| + tr\left(\Sigma_{ref}^{-1}\Delta\Sigma\right) - \frac{1}{2}tr\left(\left(\Sigma_{ref}^{-1}\Delta\Sigma\right)^2\right),$$

$$\text{and } \log\left(\frac{|\Sigma_{new}|}{|\Sigma_{ref}|}\right) \approx tr\left(\Sigma_{ref}^{-1}\Delta\Sigma\right) - \frac{1}{2}tr\left(\left(\Sigma_{ref}^{-1}\Delta\Sigma\right)^2\right).$$

This yields:

$$D_{KL}(\mathrm{N}_{new}||\mathrm{N}_{ref}) \approx \frac{1}{4}tr\left(\left(\Sigma_{ref}^{-1}\Delta\Sigma\right)^2\right).$$

(2) Relation of DISCO to KL divergence

KL divergence depends on a weighted squared sum of $\Delta\Sigma$, with weights given by $\Sigma_{ref}^{-1}$. In contrast, DISCO, focuses on the raw squared differences:

$$DISCO(\Sigma_{ref}, \Sigma_{new}) \propto \sum_{i,j} \left( [\Sigma_{ref}]_{ij} - [\Sigma_{new}]_{ij} \right)^2 = \log(||\Delta\Sigma||_F^2)$$

When principal component analysis yields $\Sigma_{ref} = I$(independent components).we have $\Sigma_{ref}^{-1} = I$ .Consequently, the KL divergence approximation simplifies to $tr\left( \left( \Sigma_{ref}^{-1} \Delta\Sigma \right)^2 \right) = ||\Delta\Sigma||_F^2$. In this case, the KL divergence is proportional to the squared Frobenius norm of $\Delta\Sigma$, which aligns with DISCO. For non-identity $\Sigma_{ref}$, the KL divergence emphasizes changes in directions where $\Sigma_{ref}$ has low variance (via $\Sigma_{ref}^{-1}$), while DISCO assigned weights with prior information. In high dimensions, $\Sigma_{ref}^{-1}$ and $|\Sigma_{ref}|$ are sensitive to noise in $\Sigma_{ref}$ estimates, leading to volatile KL values. Unlike KL divergence, DISCO avoids unstable terms like $\Sigma_{ref}^{-1}$ and $|\Sigma_{ref}|$, as its core sum $||\Delta\Sigma||_F^2$ scales robustly.

(4). Refined explanation of DISCO

DISCO quantifies the difference between a perturbed covariance matrix (after adding a new sample) and a reference covariance matrix. For a system with *m* biomarkers, DISCO is computed as the quadratic sum of differences in Pearson Correlation Coefficients (PCC) between the reference group (*n* samples) and the perturbed group (*n*+1 samples):

$$DISCO = \log(\mathrm{n}^2 * \textstyle\sum_{i=1}^{m} \sum_{j=1}^{i} w_{ij} ([C_{ref}]_{ij} - [C_{new}]_{ij})^2).$$

The $[C_{ref}]_{ij}$ is the PCC between the biomarker *i, j*, which was calculated as followed:

$$[C_{ref}]_{ij} = \frac{1}{n-1} \sum_{k=1}^{n} \left( \frac{b_{ik} - \mu_i^{ref}}{\sigma_i^{ref}} \cdot \frac{b_{jk} - \mu_j^{ref}}{\sigma_j^{ref}} \right)$$

where, $b_{ik}$, and $b_{jk}$ were values of biomarker *i*, *j* in sample *k, and* $\mu_i^{ref}$, $\sigma_i^{ref}$, $\mu_j^{ref}$ and $\sigma_j^{ref}$ were mean, variance of biomarker *i* and *j* in reference group, respectively.

As for the perturbed covariance, we added a single individual to the reference groups, respectively. Taking the sample h as an example, the elements of perturbed PCC were as followed:

$$[C_{new}]_{ij} = \frac{1}{n} \sum_{k=1}^{n+1} \left( \frac{b_{ik} - \mu_i^{ref+h}}{\sigma_i^{ref+h}} \cdot \frac{b_{jk} - \mu_j^{ref+h}}{\sigma_j^{ref+h}} \right)$$

When the size of reference group is large enough, there will be $\mu^{ref+h} \to \mu^{ref}$, $\sigma^{ref+h} \to \sigma^{ref}$, hence:

$$[C_{new}]_{ij} \approx \frac{1}{n}\sum_{k=1}^{n}\left(\frac{b_{ik}-\mu_i^{ref}}{\sigma_i^{ref}}\cdot\frac{b_{jk}-\mu_j^{ref}}{\sigma_j^{ref}}\right)+\frac{1}{n}\frac{b_{ih}-\mu_i^{ref}}{\sigma_i^{ref}}\cdot\frac{b_{jh}-\mu_j^{ref}}{\sigma_j^{ref}}.$$

This simplifies the perturbed PCC to:

$$[C_{new}]_{ij} \approx [C_{ref}]_{ij} + \frac{1}{n}\left(\frac{b_{ih}-\mu_i^{ref}}{\sigma_i^{ref}}\cdot\frac{b_{jh}-\mu_j^{ref}}{\sigma_j^{ref}} - [C_{ref}]_{ij}\right)$$

Next, the DISCO can be calculated as:

$$DISCO \approx \sum_{i=1}^{m}\sum_{j=1}^{i}\left(\frac{b_{ih}-\mu_i^{ref}}{\sigma_i^{ref}}\cdot\frac{b_{jh}-\mu_j^{ref}}{\sigma_j^{ref}} - [C_{new}]_{ij}\right)^2$$

The $n^2$ scaling factor adjusts for the reference sample size, ensuring the metric is invariant to the number of samples in the reference group. To prioritize biomarkers associated with aging, weights are applied:

$$DISCO = \mathrm{n}^2 * \sum_{i=1}^{m}\sum_{j=1}^{i} W_{i,j} * ([C_{ref}]_{ij} - [C_{new}]_{ij})^2.$$

and $W_{i,j} = \frac{PCC_{biomarker_i,age} * PCC_{biomarker_j,age}}{\sum_{i,j} PCC_{biomarker_i,age} * PCC_{biomarker_j,age}}$

The $\sum\sum PCC_{biomarker_i,age} * PCC_{biomarker_j,age}$ was the quadratic sum of PCC between biomarker *i, j*, and age, working as a scale factor.

**5. Simulations based on multivariable Gaussian distribution**

Two datasets were generated to compare the performance of Mahalanobis, DISCO, entropy and KL distances in capturing covariance structure differences, a 2-dimensional dataset and a 10-dimensional dataset. Subesquently, we used additional 10-dimensional datasets to compare the four metrics. Next, we added random Gaussian noise to the datasets and observed how the various metrics increased. Lastly, we compared the performance of these simulated survival datasets for predicting mortality when mortality is minimized near the median values.

**5.1. 2-dimensional dataset**: 1,000 samples from a bivariate normal distribution $\mu = [0, 0]^T$ and $\Sigma = [[1, \rho], [\rho, 1]]$, inducing weak (ρ= 0.1) and strong (ρ= 0.7) correlation coefficients. To visualize the distance distributions, we evaluated each metric over a 100 x 100 grid spanning 1.5 times the data range in this 2D space. Distances were normalized to the [0, 1] range (using (x-min)/(max-min)), in which the raw KL value was log-tranfromed for contour plots.

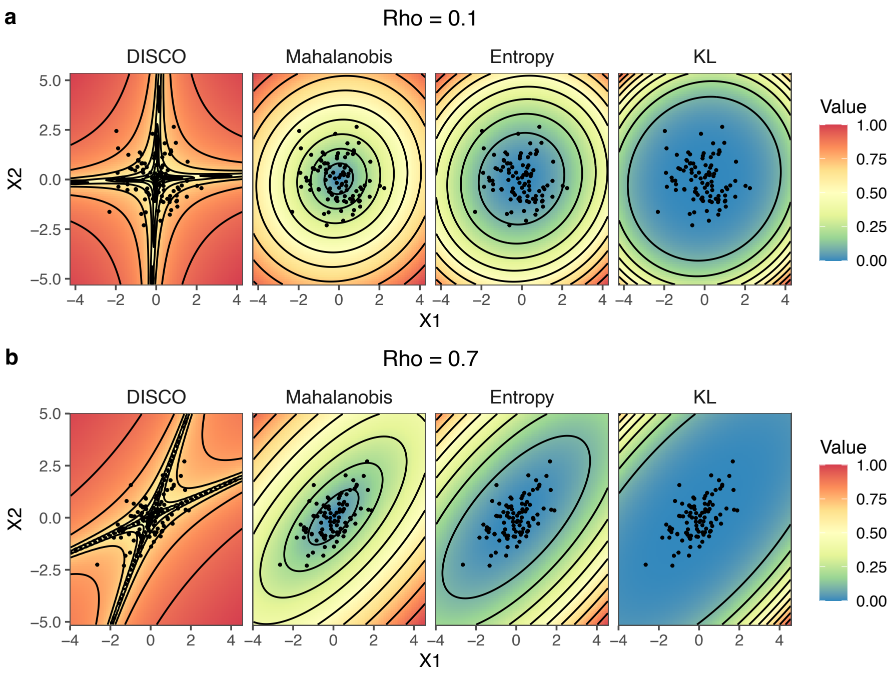


**Figure S1.** The contour distributions of the three distances across the 2D space for correlation coefficients ρ = 0.1 (a) and ρ = 0.7 (b). KL: KL-divergence distance.

**5.2. 10-dimensional datasets**: 1,000 samples were generated from a normal distribution $N(0, \Sigma)$, using a Toeplitz[1] covariance matrix (ρ= 0.1, 0.7, respectively), to create dimensionally decaying correlations. This structure mimics high-dimensional systems where adjacent features exhibit stronger dependencies. Principal Component Analysis (PCA) was applied to project data onto the PC1-PC2 subspace. A 100 x 100 grid in this subspace was back-projected to the original 10D space. Distances were computed for each grid point relative to the reference dataset's empirical covariance and normalized to [0, 1] for cross-metric comparability. For visualization: contour plots in the PC1-PC2 plane illustrated spatial patterns of each distance, overlaid with 100 randomly sampled reference points. Spearman's rank correlation (ρ) between metric pairs, were supplemented by scatter plots.

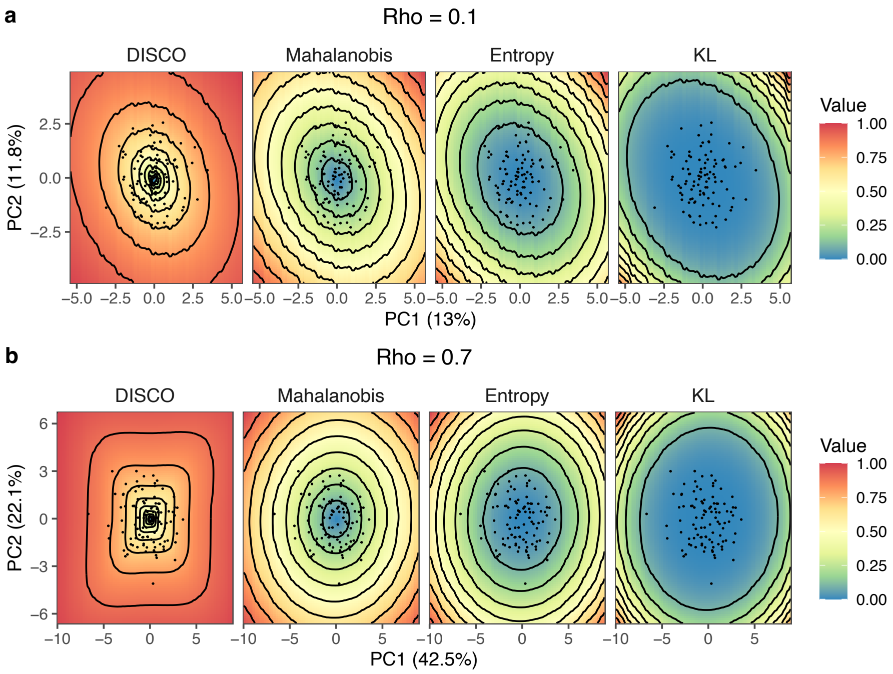


**Figure S2.** The contour distributions of the three distances in the PC1-PC2 plane for the 10D datasets with Toeplitz matrix Σ (ρ = 0.1, ρ = 0.7).

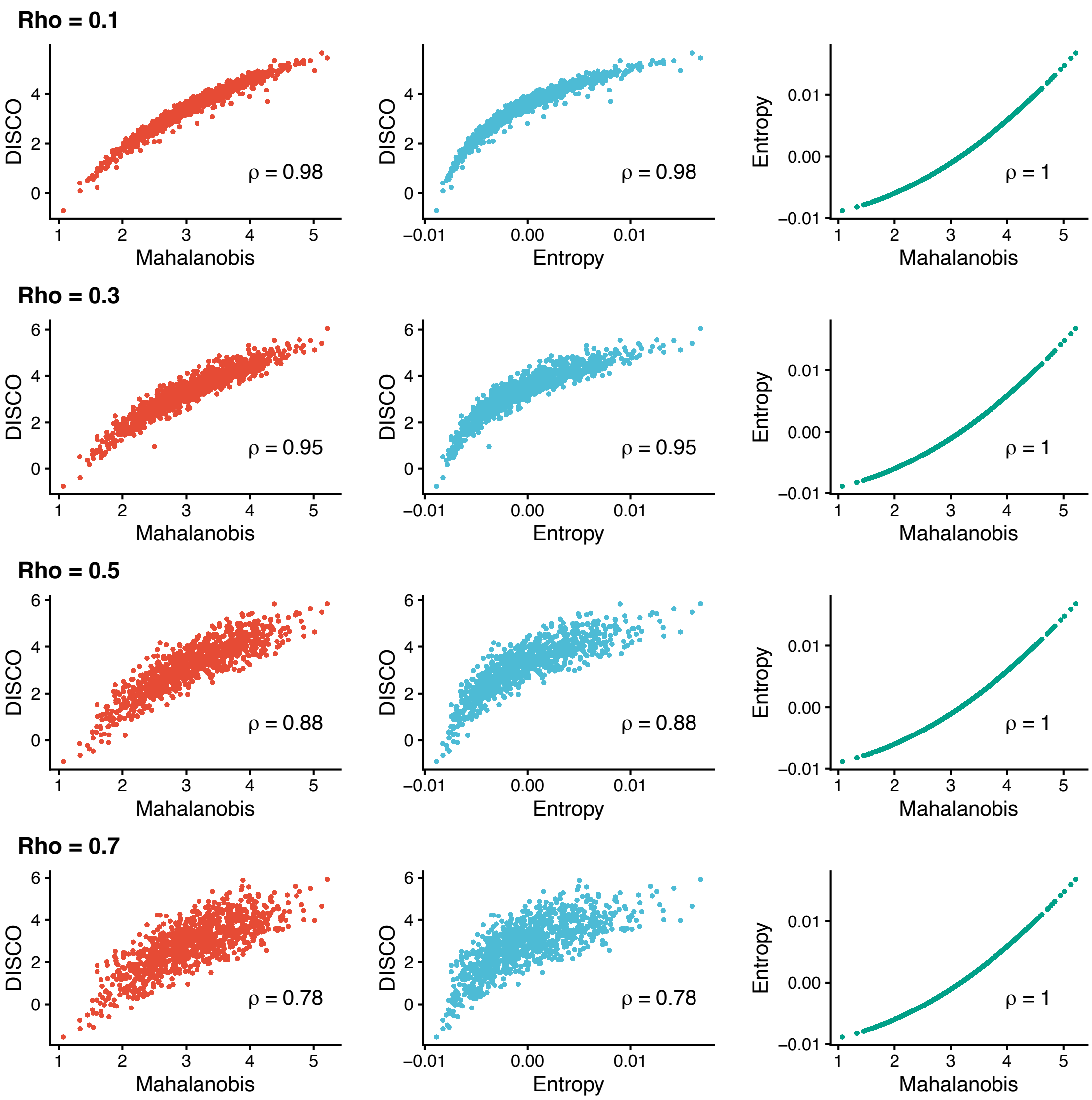


**Figure S3.** The pairwise correlations (Spearman ρ) among DISCO, Mahalanobis, and Entropy distances based on the 10D generated data with Toeplitz matrix Σ (ρ = 0.1, 0.3, 0.5, 0.7).

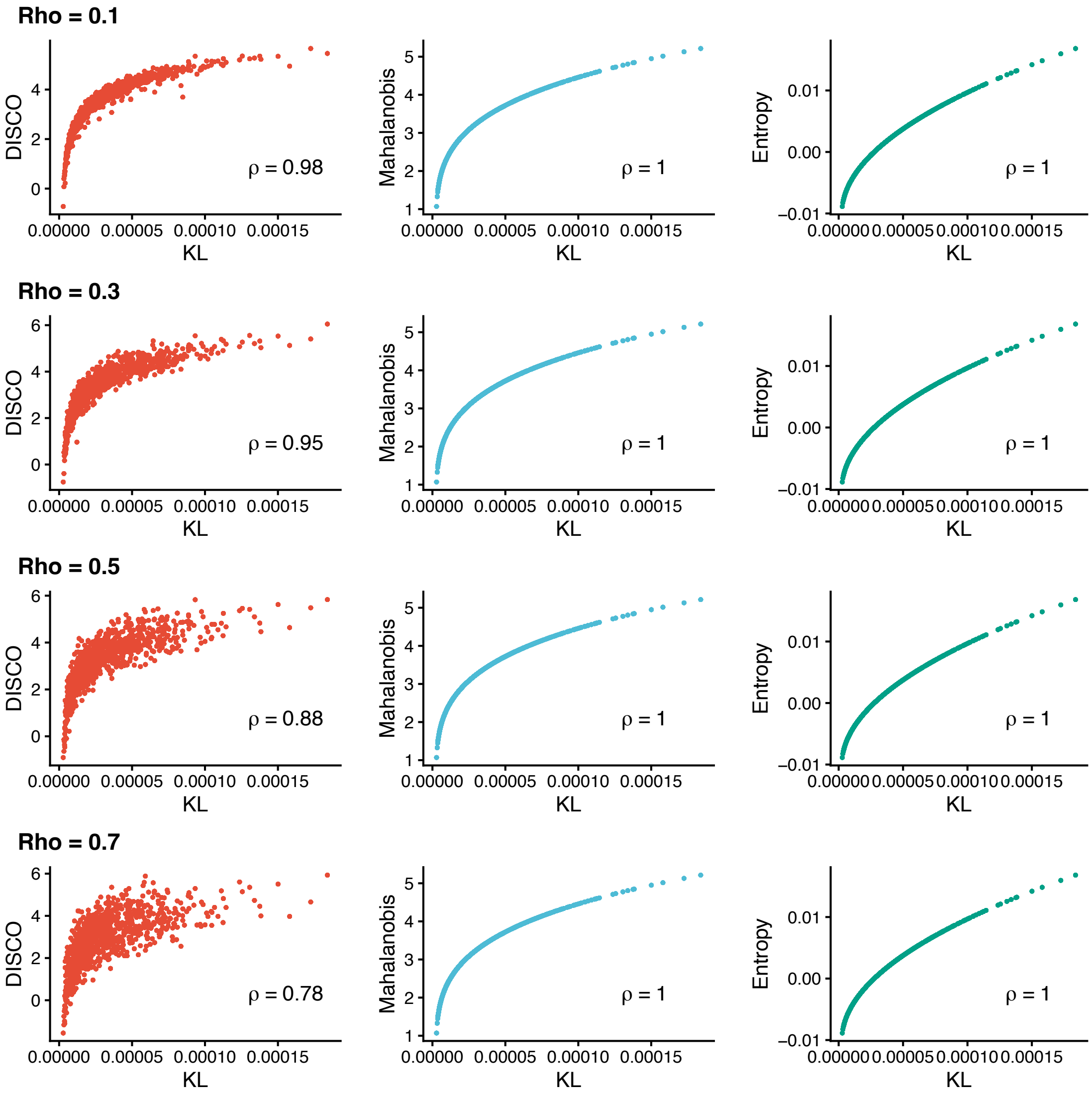


**Figure S4.** The pairwise correlations (Spearman ρ) among DISCO, Mahalanobis and KL divergence. Top row: ρ = 0.1; 2nd row: ρ = 0.3; 3rd row: ρ = 0.5; bottom row: ρ = 0.7.

**5.3. Associations with KL divergence**. Using 10-dimensional datasets, we evaluated three distance measures against KL divergence under different covariance conditions. KL divergence was quantified before adding the additional individual data into the reference dataset. All measures were strongly correlated with KL divergence when variables were nearly independent (ρ = 0.1). As covariance increased, these correlations declined for the Mahalanobis and entropy distances but were maintained for DISCO.

**5.4. Addition of noise**. We further tested the measures by introducing entropy through the addition of independent Gaussian noise to each variable in the dataset. This process weakens the existing correlation structure, simulating a biological scenario where a process perturbs individual variables independently. As expected, all measures increased with the added noise. The fact that DISCO increases in response to this entropy-inducing manipulation validates it as a robust measure of entropy.

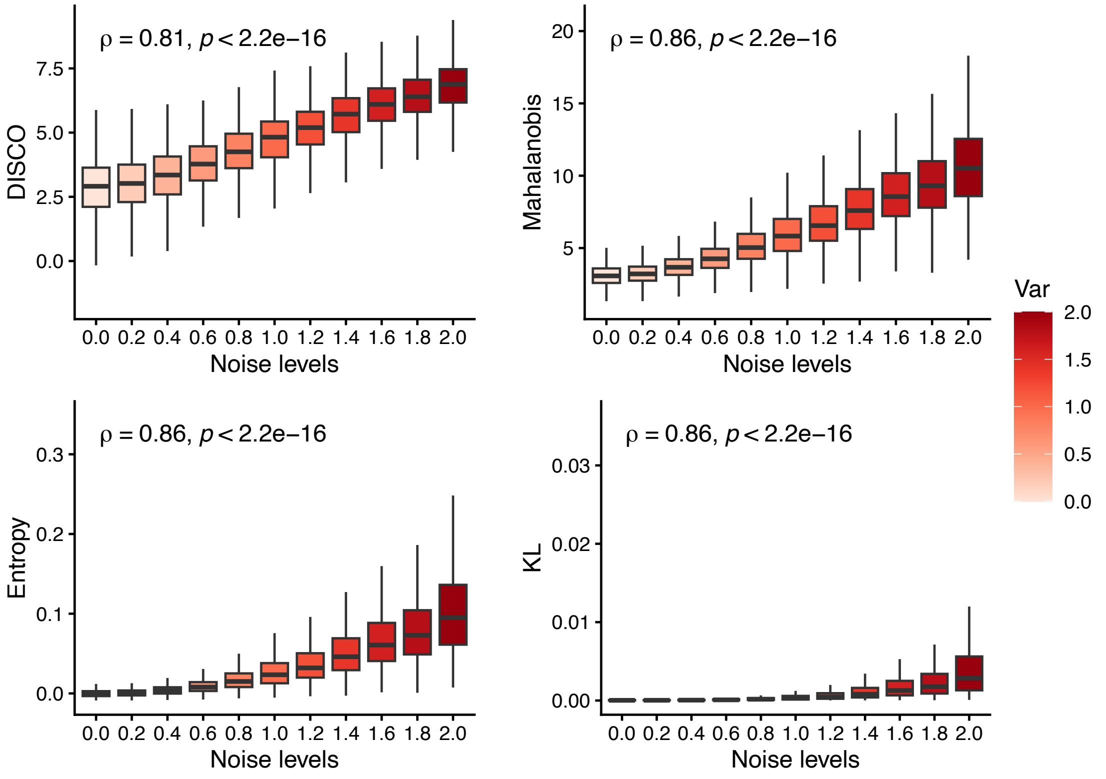


**Figure S5. Increase in entropy measures as a function of Gaussian noise level (σ).** The noise ϵ was sampled from a normal distribution N(0, σ²).

**5.5 Benchmarking three distance measures**. To evaluate how well three distance measures capture the risk of adverse outcomes, we generated simulation datasets using four survival distributions: Cox proportional hazards, Exponential, Gompertz, and Weibull. The hazard functions were defined as:

$$\text{Cox: } h(t, X) = h_0(\text{t}) * \exp{(Z)}$$

$$\text{Exponential: } h(t, X) = \lambda * \exp{(Z)}$$

$$\text{Gompertz: } h(t, X) = \lambda e^{\gamma t} * \exp{(Z)}$$

$$\text{Weibull: } h(t, X) = \lambda \gamma t^{\gamma - 1} * \exp{(Z)},$$

where λ is the scale parameter and γ is the shape parameter.

The predictor Z was modeled as a quadratic function of covariates X: $Z = X^2\beta + \epsilon$, with $\epsilon \sim N(0, \sigma^2)$. This functional form reflects the hypothesis that risk is minimized near the median biomarker level (X=0) and increases with greater deviation.

We simulated six scenarios by varying the number of covariates (10, 50, 100) under two levels of Gaussian noise ($\sigma$=0.2, 1). Each scenario generated 1000 samples (X) from a normal distribution $N(0, \Sigma)$, with a Toeplitz covariance matrix (ρ= 0.1, 0.3, 0.5, 0.7, respectively), using the R 'mvtnorm' package. Survival times and event statuses were then generated for each distribution using the R 'simsurv' and 'coxed' packages, with parameters set as: Exponential ($\lambda = 0.1$), Gompertz ($\lambda = 0.1, \gamma = 0.1$), Weibull ($\lambda = 0.1, \gamma = 0.1$), Cox (NULL for $\lambda, \gamma$).

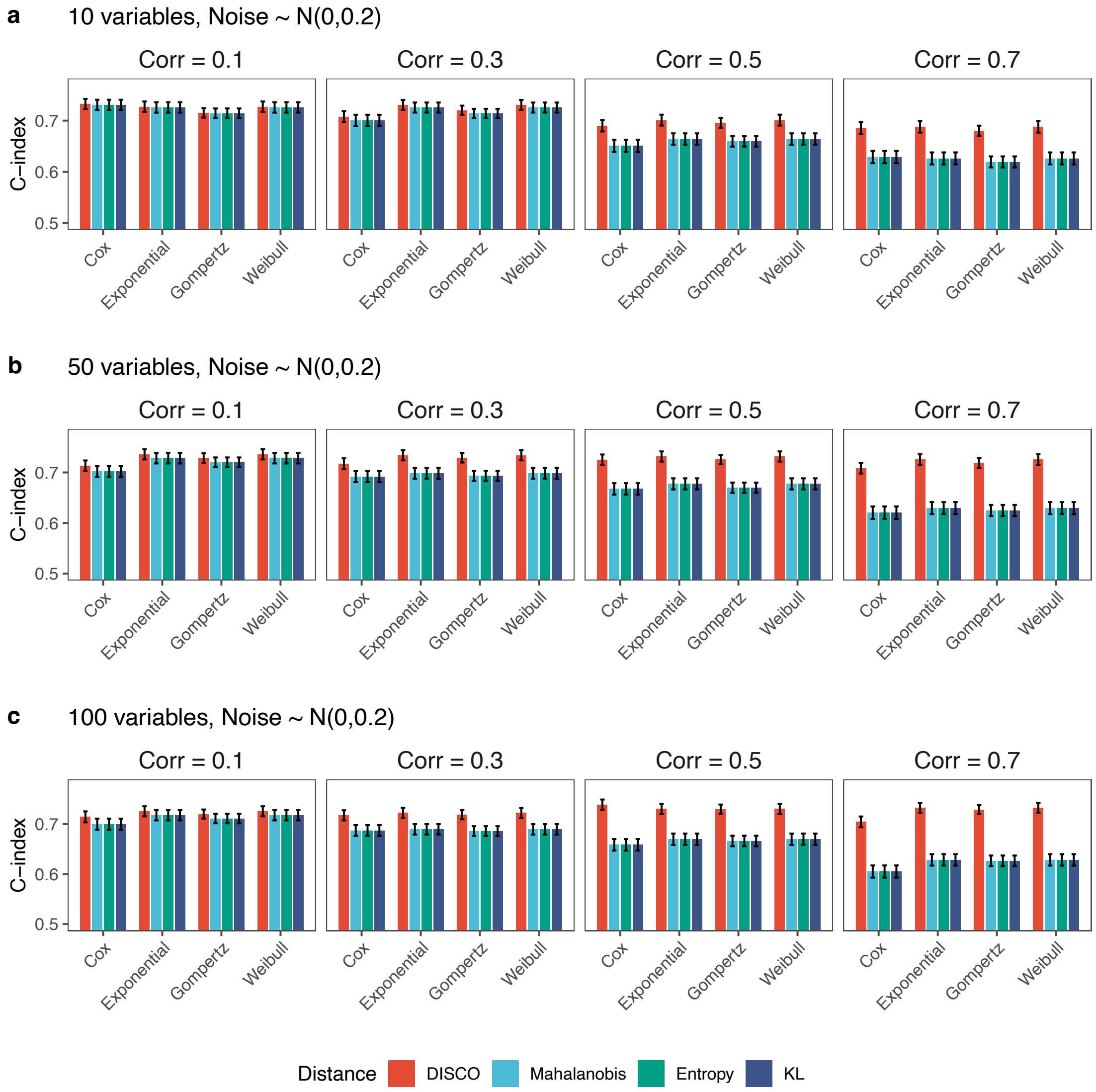


**Figure S6. Concordance of survival analysis for three distances.** The noise item ϵ was sampled from a normal distribution N(0, $\sigma^2$), and σ = 0.2. The covariates (n = 10, 50, 100)were generated from Gaussian distributions with Toeplitz matrix Σ (ρ = 0.1, 0.3, 0.5, 0.7).

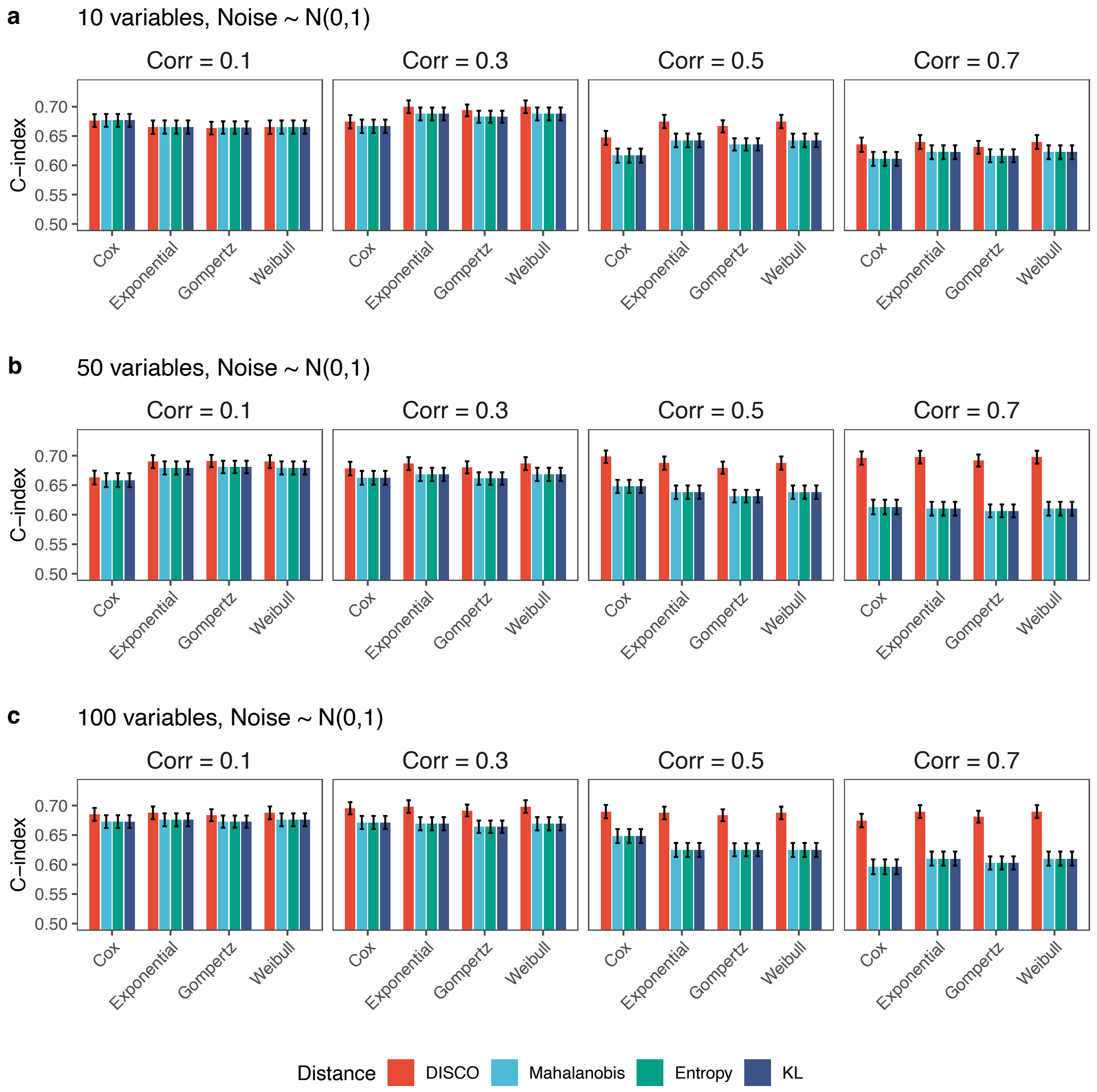


**Figure S7. Concordance of survival analysis for three distances.** The noise item ϵ was sampled from a normal distribution N(0, $\sigma^2$), and $\sigma = 1$. The covriates (n = 10, 50, 100)were generated from Guassian distributions with Toeplitz matrix Σ (ρ = 0.1, 0.3, 0.5, 0.7).

**5.6. Interpretation of simulations**. The distributional simulations (Figs S1-S2) show that, compred to $d_M$, $d_E$ and $d_{KL}$, DISCO ($d_d$) detects finer-scale differences at the low end of the scale. This is also apparent in Fig. S3, where the functional forms apparent in the scatter plots show that $d_M$, $d_E$ and $d_{KL}$ fail to detect meaningful differences at low divergence, in contrast to $d_d$. Whether this is advantageous or disadvantageous for $d_d$ as a measure of entropy in the context of aging and health depends on whether those finer-scale differences are meaningful in terms of health status; results in the main text suggest they are, at least in higher-dimensional datasets.

When Gaussian noise is added to the dataset, all three entropy measures (as well as KL divergence) show monotonic increases, as expected. In the case of DISCO, the variance does not increase meaningfully with the noise, whereas for the other three metrics, there is lower variance at low noise and higher variance at high noise. Spearman correlations with noise levels are of comparable magnitudes across metrics.

When mortality simulations are added to benchmark performance, little difference between the metrics is observed when the correlation structure is weak ($\rho = 0.1$), but as the correlations become stronger, DISCO starts to outperform the other three metrics (**Figs S6-7**). This conclusion holds regardless of the number of variables or the quantity of noise used in the simulations. It should be noted that these simulations assume minimum mortality at the median of the biomarker values.

Globally, these simulations suggest that all four entropy measures are theoretically valid and could work in practice. DISCO is expected to perform slightly better under conditions where there is important signal at relatively low levels of entropy/dysregulation, and when there are strong correlations among the variables. There are no situations in which other metrics meaningfully outperform DISCO.
